# GENOME-WIDE ASSOCIATIONS OF AORTIC DISTENSIBILITY SUGGEST CAUSAL RELATIONSHIPS WITH AORTIC ANEURYSMS AND BRAIN WHITE MATTER HYPERINTENSITIES

**DOI:** 10.1101/2021.09.01.21262202

**Authors:** Catherine M Francis, Matthias E Futschik, Jian Huang, Wenjia Bai, Muralidharan Sargurupremraj, Enrico Petretto, Amanda SR Ho, Philippe Amouyel, Stefan T Engelter, James S Ware, Stephanie Debette, Paul Elliott, Abbas Dehghan, Paul M Matthews

## Abstract

Aortic dimensions and distensibility are key risk factors for aortic aneurysms and dissections, as well as for other cardiovascular and cerebrovascular diseases. We tested genome-wide associations of ascending and descending aortic distensibility and area derived from cardiac magnetic resonance imaging (MRI) data of up to 32,590 Caucasian individuals in UK Biobank. We identified 102 loci (including 31 novel associations) tagging genes related to cardiovascular development, extracellular matrix production, smooth muscle cell contraction and heritable aortic diseases. Functional analyses highlighted four signalling pathways associated with aortic distensibility (TGF-β, IGF, VEGF and PDGF). We identified distinct sex-specific associations with aortic traits. We developed co-expression networks associated with aortic traits and applied phenome-wide Mendelian randomization (MR-PheWAS), generating evidence for a causal role for aortic distensibility in development of aortic aneurysms. Multivariable MR suggested a causal relationship between aortic distensibility and cerebral white matter hyperintensities, mechanistically linking aortic traits and brain small vessel disease.

## INTRODUCTION

The aorta acts as both conduit and buffer^1^, conveying oxygenated blood from the heart to the systemic circulation, and dampening the pulse pressure to which peripheral circulations are subjected. Diseases affecting the aorta are common and their complications are associated with a high mortality even in young people. Quantitative aortic traits (aortic dimensions and functional measures) can predict progression of these aortopathies. For example, the elastic function of the thoracic aorta and aortic dimensions are key determinants of rates of growth of thoracic aortic aneurysms^2-4^. At a population level, aortic traits are also clinically important predictors of risks of cardiovascular and cerebrovascular diseases^5-10^.

The decline in elastic function with age may be measured as a decrease in distensibility, a factor independently predictive of cerebral microvascular disease, the development of age-related dementia and neurodegenerative changes of Alzheimer’s Disease (AD)^11-13^. Recent data also have provided evidence for an association of aortic distensibility with cognitive performance in the general population^14^. White matter hyperintensities (WMH) represent the most common brain imaging feature of small vessel disease and predict mortality and morbidity with aging (including risks of stroke (ischemic and hemorrhagic), dementia, and functional impairment^15-18^). Aortic stiffness is a stronger predictor of WMH volume than blood pressure or hypertension alone^19, 20^ and has effects additive to those of hypertension in predicting WMH^19-21.^ The genomic bases of these relationships have not been well explored to date.

With age, aortic stiffening arises from changes in composition of the aortic wall, including degradation of the elastic fibres and decreased cellularity, along with a relative increase in the collagen content of the aorta (although the absolute amount decreases)^22, 23^. In addition, extracellular matrix proteins themselves undergo conformational and biochemical changes which alter their passive mechanical properties. These remodelling processes are driven by TGF-β signalling pathways and accelerated by oxidative stress and inflammation^24^ acting on the cells in the aortic wall. These cellular and molecular drivers of worsening aortic elastic function are reflected in macroscopic changes with age.

Here, we used convolutional neural networks for automated aortic segmentation^25^ to measure ascending and descending aortic areas and distensibilities on cardiac magnetic resonance (MRI) images from UK Biobank, which is currently the largest cardiac imaging epidemiological study^26^. We have described our approach to derivation of imaging derived quantitative aortic traits and the distribution of these traits in a smaller group from the same population in an earlier report^25, 27^. We derived six aortic traits (ascending aortic distensibility (AAdis), descending aortic distensibility (DAdis), maximum ascending aortic area (AAmax), minimum ascending aortic area (AAmin), maximum descending aortic area (DAmax) and minimum descending aortic area (DAmin)) in up to 32,590 (depending on the specific trait) UK Biobank participants, who were free from known aortic disease We then performed a genome-wide association study (GWAS) of the six CMR-derived aortic traits and carried out functional analyses and a series of Mendelian Randomization studies to investigate possible causal associations of the aortic measures with aortic aneurysms and brain small vessel disease. We also explored the bidirectional relationship of aortic traits with indices of blood pressure.

## RESULTS

Study cohort exclusions are presented in Supplementary Figure S1. The distributions of the aortic traits are shown in Supplementary Figure S2.

### SNP-based heritability

We estimated the proportion of the variability in aortic traits that could be attributed to common genetic variation from an analysis of SNP-based heritability (*h*^2^_SNP_) using linkage disequilibrium score regression (LDSC). *h*^2^_SNP_ estimates ranged from 0.10 (for DAdis single trait) to 0.41 (for AAmax). We also tested for heritability of distensibility traits using multi-trait analysis (MTAG, *h*^2^_SNP_=0.21 for DAdis and *h*^2^_SNP_=0.24 for AAdis).

### Phenotypic and genotypic correlations between traits

We found strong phenotypic and genotypic correlations between maximum and minimum aortic areas (phenotypic r=0.99, p<2.2×10^−16^; genotypic r_g_=0.99, p<1×10^−50^ for the ascending aorta; phenotypic r=0.98, p<2.2×10^−16^; genotypic r_g_ =0.99, p<1×10^−50^ for the descending aorta).There were lower correlations between ascending and descending aortic traits (phenotypic r=0.60, p<2.2×10^−16^ and genotypic r_g_=0.45, p<1.7×10^−25^ for the minimum aortic areas and phenotypic r=0.74, p<2.2×10^−16^ and genotypic r_g_=0.45, p<9.25×10^−7^ for distensibilities) consistent with known biological and functional differences along the course of the aorta^1^. Correlations are presented in Supplementary Figures S3 and S4.

### Single and multi-trait aortic GWAS

Our stage 1 GWAS (N=32,590 for areas and N=29,895 for distensibility) identified a total of 95 significant loci (using a genome-wide significance threshold of p<5×10^−8^) across the six traits, 94 of which are autosomal with one localised to the X chromosome. Genomic inflation was within acceptable limits for all traits (λ=1.147 for the area traits; λ=1.047 for the distensibility traits). We took advantage of the correlation between the aortic traits to enhance power for discovery of loci by performing multi-trait analysis (MTAG)^28^ as a second stage of our GWAS. Use of MTAG combining all six phenotypes increased the number of significant loci for the distensibility traits from 10 to 26 for the ascending aorta and from 7 to 13 for the descending aorta (Table 1), and the total number of significant loci across all aortic traits to 102. Figure 1A shows the Manhattan and QQ plots from GWAS of ascending and descending aortic minimum areas, which overlap almost completely with the findings for the corresponding maximum areas. Figure 1B shows the results of the MTAG analysis of ascending and descending aortic distensibilities. GWAS summary statistics from 9,753,033 variants with a minor allele frequency (MAF) ≥ 0.01 for the stage 1 and stage 2 (MTAG) analyses are shown in Supplementary Figures S5-S8.

**TABLE 1:**
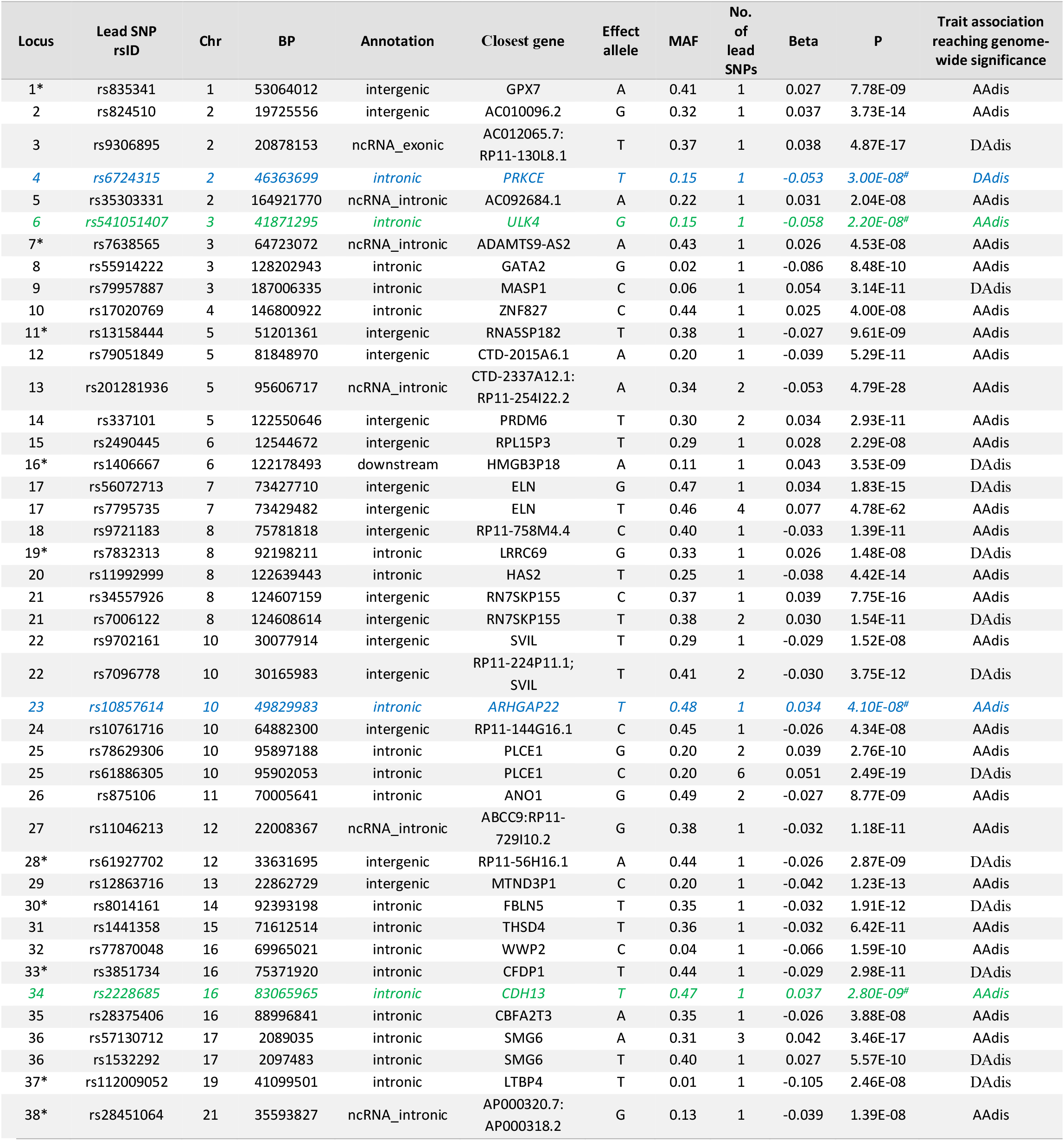
Significant associations with distensibility in single and multi-trait genome-wide analyses. Summary statistics are shown for lead SNPs which were genome-wide significant (p<5×10^−8^) in MTAG analysis (loci in black) apart from the four loci (in green and blue) which were significant in single trait analysis only. Where loci overlapped, different lead SNPs are listed separately under the same locus number. Green = genome-wide significant in only the single trait analysis, not the multi-trait analysis, but also significantly associated with aortic area traits. MAF=Minor Allele Frequency; Chr=chromosome, BP= position (GRCh37). Blue = genome-wide significant in only the single trait analysis of distensibility and in no other aortic traits. *=novel locus not identified in previous GWAS of ascending or descending aortic traits.

**FIGURE 1A:**
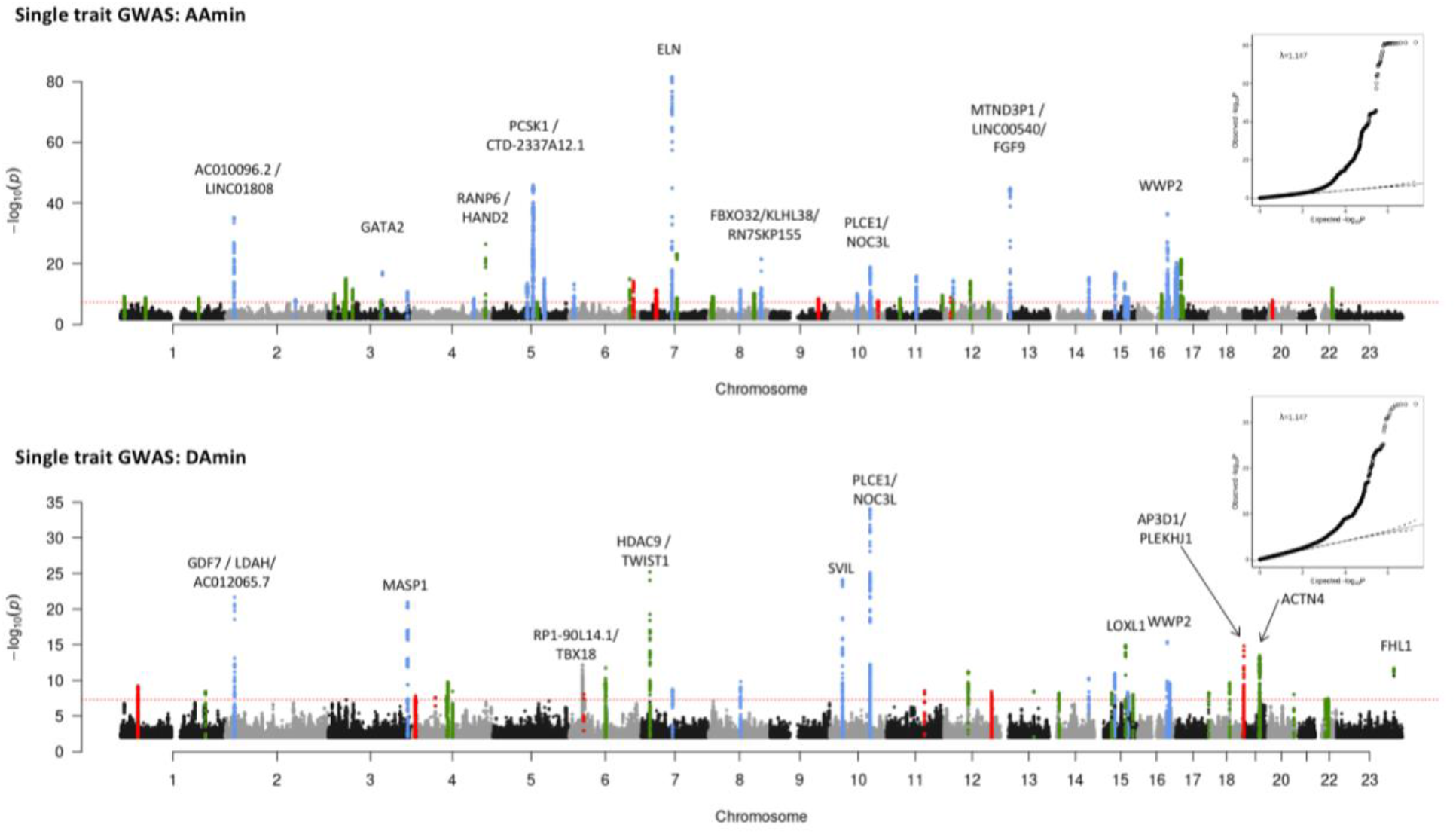
Manhattan plots of summary statistics from single-trait GWAS of aortic areas. Red dashed line shows the genome-wide significance threshold of *P* = 5 × 10^−8^. Genomic inflation (*λ*) = 1.147 (AAmin and DAmin). Annotations of selected loci show the nearest gene and additional manual annotation of likely candidate gene(s) at the locus where appropriate. Blue: locus is genome-wide significant in multiple aortic traits, green: locus is genome wide significant only in the corresponding trait and with nominal significance (p<0.01) in other traits, red: genome-wide significant only in the corresponding trait without even nominal significance in other traits. QQ plots are shown as inserts in corresponding panels.

**FIGURE 1B:**
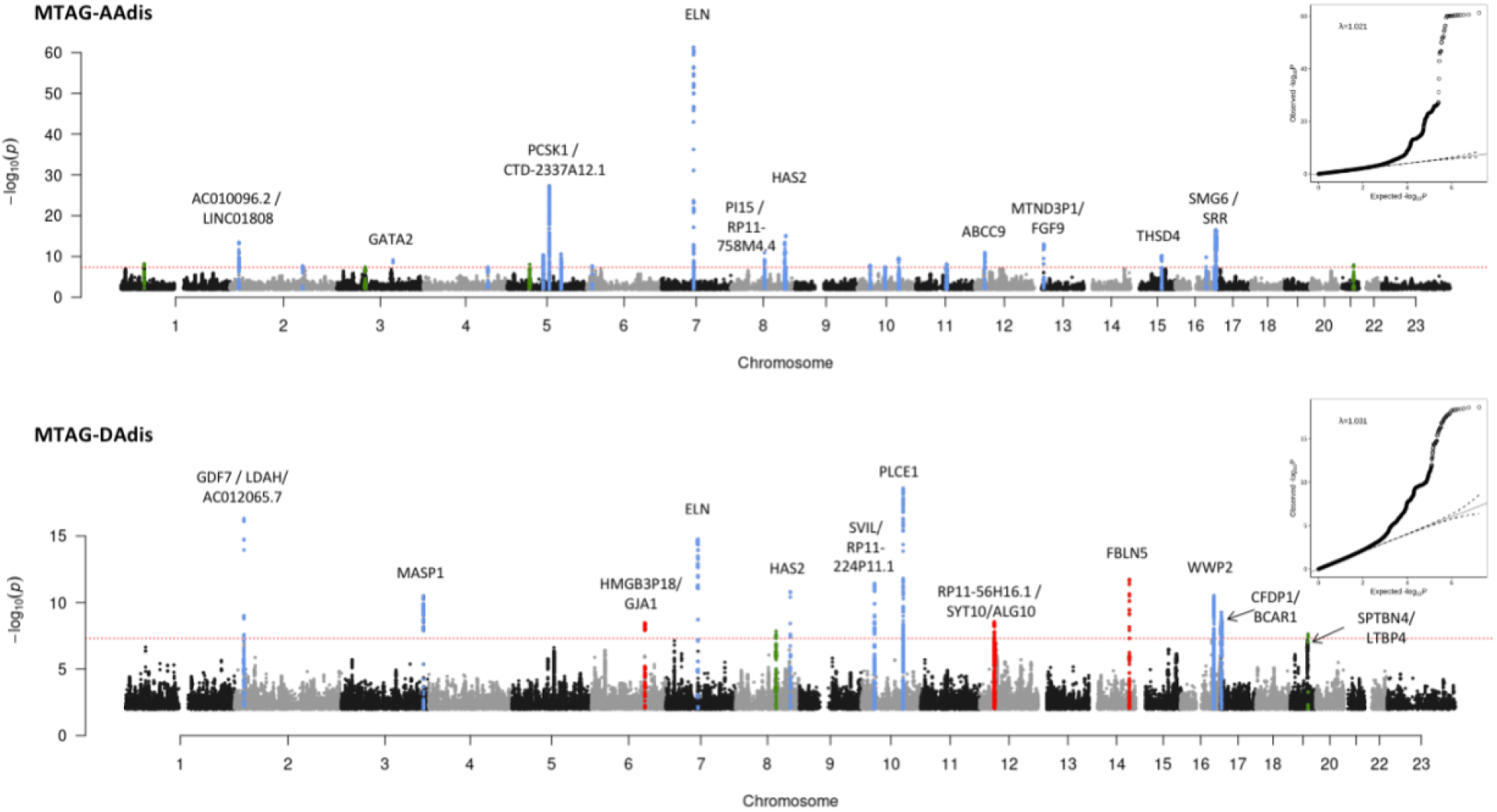
Manhattan plots of summary statistics from multi-trait GWAS (MTAG) of aortic distensibility. All six traits were used for the MTAG analysis. Twenty-six association signals were identified in multi-trait analysis of AAdis and thirteen in multi-trait analysis of DAdis. Genomic inflation (*λ*) = 1.021 (AAdis) and 1.031 (DAdis). Red dashed line shows the genome-wide significance threshold of *P* = 5 × 10^−8^. Blue: locus is genome-wide significant in multiple aortic traits, green: locus is genome wide significant only in the corresponding trait and with nominal significance (p<0.01) in other traits, red: genome-wide significant only in the corresponding trait without even nominal significance in other traits. QQ plots are shown as inserts in corresponding panels.

Individual loci were annotated with *cis*-expression quantitative trait loci (eQTL) and splice quantitative trait loci (sQTL) data from GTEx v8^29^. Twenty-four of the 38 loci associated with distensibilities had lead SNPs which were significant eQTLs or sQTLs for nearby genes in arterial tissue.

The most significant associations with ascending aortic distensibility were: rs7795735, 12.6 kilobases upstream of *ELN*, a gene encoding elastin; rs201281936, which is in a lncRNA (*CTD-2337A12*.*1)* 119 kilobases 3’ of *PCSK1* (Proprotein Convertase Subtilisin/Kexin Type 1); rs57130712, which is in a locus spanning *SMG6 (*SMG6 Nonsense Mediated MRNA Decay Factor) and *SRR* (Serine Racemase), and which is the most significant eQTL for SRR in arterial tissue (p=2×10^−20^, normalised effect size (NES)= -0.39)^29^; and rs34557926, an intronic variant in *HAS2* (Hyaluronan Synthase 2).

The strongest associations with descending aortic distensibility were different. The most significant association was with rs61886305, an intronic variant in *PLCE1 (*Phospholipase C Epsilon 1). This lead SNP is a strong eQTL for *PLCE1* in arterial tissue (p=1.1×10^−8^, NES= -0.20)^29^. The next strongest association was with rs9306895, an intronic variant in *GDF7* (Growth Differentiation Factor 7), which is a strong eQTL for both *GDF7* and *LDAH* (Lipid Droplet Associated Hydrolase) in aorta^29^ *(GDF7*: p=3.6 ×10^−9^, NES= 0.20; *LDAH*: p=7.1×10^−28^, NES= 0.53). A locus spanning *ELN* was associated with DAdis, with the lead SNP less than 1.8kb away from the lead SNP for AAdis at this locus, and in strong LD with it (R^2^=0.96; D’=1). A locus in *FBLN5* (lead SNP rs8014161) was associated with descending, but not ascending distensibility.

There were four loci associated at genome-wide significance with aortic distensibilities which lost genome-wide significance in the MTAG analysis (three associated with AAdis and one with DAdis; see Table 1 for details). Two of these were not significantly associated with any other aortic traits: rs6724315 in *PRKCE* and rs10857614 in *ARHGAP22*. The latter is a strong eQTL for ARHGAP22 in aorta (p=2.6×10^−46^, NES= 0.58), providing additional evidence for its biological relevance.

We compared our association results for aortic areas with those reported for aortic diameters in recent preprints^30, 31^ based on the same UK Biobank imaging dataset, but using different methods and metrics for aortic dimensions. We replicated 73 of the previously reported genome-wide significant association loci and added associations for two clinically relevant phenotypes (ascending and descending aortic distensibility) to identify a further 31 novel associations. Only four of the previously reported SNPs did not replicate. Inspection of the loci for SNPs that were significant in the analysis by Pirruccello et al^30^, but not in our own, generally showed SNP p-value signals near the genome-wide significance threshold (p<5×10^−8^) in our analysis. The small differences between associations in the two studies could arise from differences in the methods used to generate quantitative phenotypes or from the differences in participant exclusions between the two studies (for example, we excluded data from participants with known diagnoses of aortic disease and those who were extreme phenotypic outliers).

Novel aortic loci associated with AAdis included lead SNPs rs7638565 near *ADAMTS9*, which is a significant eQTL for this gene in aorta (p=5.7×10^−7^, NES= -0.22) and rs835341, intronic in *GPX7* and a strong eQTL in the aorta (p=3×10^−95^, NES= -0.92). A novel locus associated with descending aortic distensibility included lead SNP rs112009052, an sQTL for *LTBP4* in fibroblasts (p=9.6×10^−67^, NES= 2.9) but not in aortic tissues that has relevance here due to this gene’s role in the TGF-β pathway.

We identified 21 novel associations with aortic areas including lead SNPs in *KALRN* and *COL21A1* associated with ascending aortic areas and SNPs at loci tagging *AFAP1, FGF5/BMP3, NOX4, FES* and *GATA5/LAMA5* associated with descending aortic areas (see Supplementary Figures S5).

### Sex-specific aortic trait GWAS analyses

We undertook sex-specific GWAS analysis of the area phenotypes. We did not perform these analyses for distensibility phenotypes due to lack of power with the smaller cohort sizes. We contrasted associations discovered for the men and women (numbers of whom were well-balanced in the cohort) using a z test. There were 18 loci (ten for AAmin and eight for DAmin) at which the differences between sexes were significant (adjusted p <0.05). Seven of these associated loci were not significant even at p<0.05 for one sex, despite reaching genome-wide significance (p<5×10^−8^) for the other (see Table 2). Amongst these sex-specific loci were rs28699256, a missense variant in in *ADAMTS7* associated with ascending aortic area in females, but not males (see Table 2; z-test for sex difference, p=7.6×10^−4^). This variant in *ADAMTS7* is in strong LD with the lead SNP associated with AAmin in the full cohort at genome-wide significance (rs7182642; R^2^=0.76, D’=0.94). Amongst the other sex-specific signals were four others with functional data supporting potential biological roles in the aorta, all of which were significant only in males: rs72765298 in *SCAI*, a strong eQTL in aorta (see Table 2; p value for eQTL in aorta=8.9×10^−18^, NES= -0.48); rs632650, a significant eQTL for *ALDH2* in aorta (see Table 2; p value for eQTL in aorta=3.3×10^−12^, NES= 0.26); rs6573268, associated with DAdis, in *CCDC175* which is a significant eQTL for this gene and others in the aorta (see Table 2; p value for eQTLs in aorta: for *CCDC175* p=1.2×10^−6^, NES = 0.35; for *RTN1* p=2.4×10^−7^, NES= 0.33 and for *L3HYPDH*, p=1.1×10^−4^, NES= 0.25); and rs35346340 in *FES*, a strong eQTL for this gene in aorta (see Table 2; p value for eQTL in aorta 2.4 × 10^−15^, NES= 0.3).

**TABLE 2:** Lead SNPs at sex-specific loci. SNPs are shown if they reach genome-wide significance (p<5×10^−8^) in one sex and are not significant (p>0.05) in the other. Gene = nearest gene. Trait = aortic trait with which association in one sex is genome-wide significant, pval = p value from sex-specific GWAS, beta= effect size from sex-specific GWAS, SE= standard error from sex-specific GWAS, sig = which sex the SNP has reached genome wide significance in, z stat = z statistic for comparison between sexes, z.pval= p value of sex comparison.

| SNP | Gene | Trait | Effect allele | Males |  |  | Females |  |  | Comparison |  |  |
| --- | --- | --- | --- | --- | --- | --- | --- | --- | --- | --- | --- | --- |
|  |  |  |  | pval | beta | SE | pval | beta | SE | sig | z stat | z.pval |
| rs72765298 | SCAI | AAmin | T | 7.10E-09 | 15.54 | 2.68 | 0.43 | 1.77 | 2.25 | M | 5.58 | 2.43E-08 |
| rs28699256 | ADAMTS7 | AAmin | T | 0.066 | -3.32 | 1.81 | 4.10E-09 | -8.92 | 1.52 | F | 3.37 | 7.63E-04 |
| rs632650 | ACAD10 | DAmin | G | 1.30E-09 | -6.76 | 1.11 | 0.54 | -0.52 | 0.85 | M | -6.33 | 2.39E-10 |
| rs6573268 | CCDC175 | DAmin | G | 1.10E-08 | 5.15 | 0.90 | 0.29 | 0.72 | 0.69 | M | 5.53 | 3.22E-08 |
| rs35346340 | FES | DAmin | G | 8.10E-09 | 4.93 | 0.86 | 0.081 | 1.14 | 0.65 | M | 5.00 | 5.62E-07 |
| rs9449999 | TBX18 | DAmin | A | 2.70E-08 | -4.52 | 0.81 | 0.12 | -0.96 | 0.62 | M | -4.94 | 7.64E-07 |
| rs577351796 | TBC1D12 | DAmin | C | 0.11 | -6.50 | 4.08 | 6.00E-11 | -19.90 | 3.04 | F | 3.74 | 1.83E-04 |

An association with rs12663193 (intronic in *ESR1*) reached genome-wide significance only in females. However, the sex difference was not significant (z-test p value = 0.06).

### Gene-based analysis and tissue specificity

We prioritized potentially causal genes at significant loci using two complementary strategies: FUMA^32^, which integrates positional mapping, eQTL associations and HiC-derived 3D chromatin interactions (see Methods) and MAGMA^33^, which aggregates SNP associations within genes. In total, FUMA identified 973 candidate genes across the six phenotypes, including 390 protein-coding genes, 164 pseudogenes, 129 lincRNAs, 115 antisense RNAs and 46 miRNAs (Supplementary Figure S9). MAGMA identified 391 candidate genes with an FDR < 0.01 (Supplementary Figure S10).The most significant gene associations (MAGMA) for ascending and descending aortic distensibilities are shown in Table 3.

**TABLE 3:**
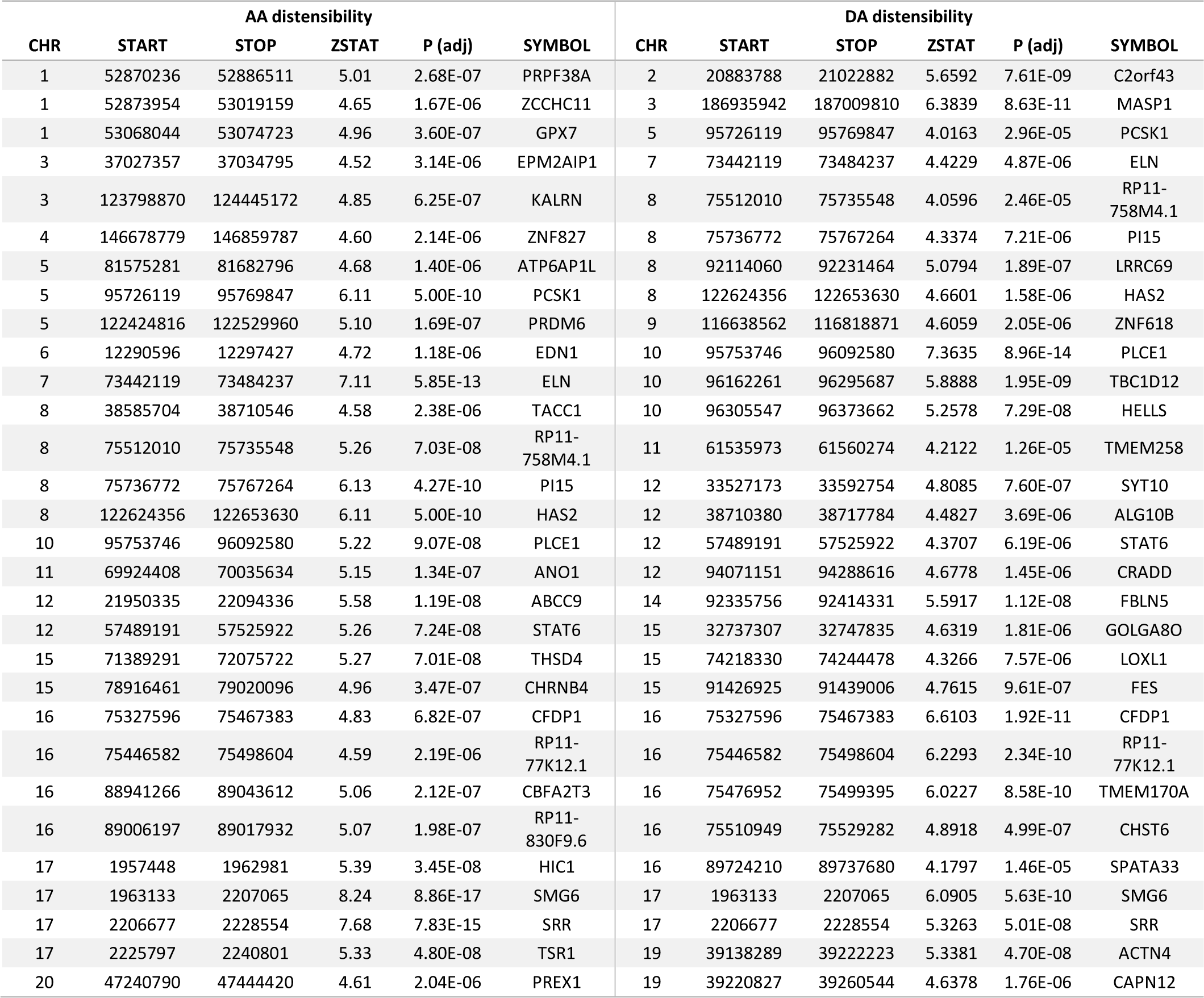
Most significant 30 genes associated with ascending and descending aortic distensibility (MAGMA). P values are converted to adjusted p-values (FDRs) using the Benjamini-Hochberg procedure.

Four genes (*MASP1, PI15, PLCE, TBC1D12* [the last likely tagging the *PLCE1* locus]) reached significance for all 6 aortic traits, with *ELN* at genome-wide significance in all traits except for DAmax, where it was just below the genome-wide significance threshold.

Tissue specificity analysis in MAGMA for genes associated with each phenotype demonstrated that these were significantly enriched for expression in aorta and in coronary artery (p value for enrichment < 1×10^−3^ in all traits), supporting the validity of our results. See Supplementary Figure S11 for further details.

### Gene set enrichment and pathways analyses

The GO terms identified by MAGMA (Supplementary Figure S12) that were most significantly associated with our aortic phenotypes highlighted processes important for the development of aortic aneurysms and dissection, such as “extracellular matrix structural constituent” and “smooth muscle contraction”, as well as GO terms related to cardiovascular development.

DEPICT implicated similar ontologies and identified three molecular pathways significantly enriched in our data (FDR<0.01) for at least one aortic trait (see Figure 2) and of nominal significance in all other traits: regulation of TGF-β signalling (AAdis nominal p value=2.76×10^−5^; FDR <0.01), IGF binding (AAdis nominal p value 1.47×10^−4^; FDR <0.01) and PDGF binding (AAdis nominal p value 7.19×10^−5^; FDR <0.01). VEGF signalling was significantly associated with ascending aortic distensibility (AAdis nominal p value 5.15×10^−5^ ; FDR <0.01).

**FIGURE 2:**
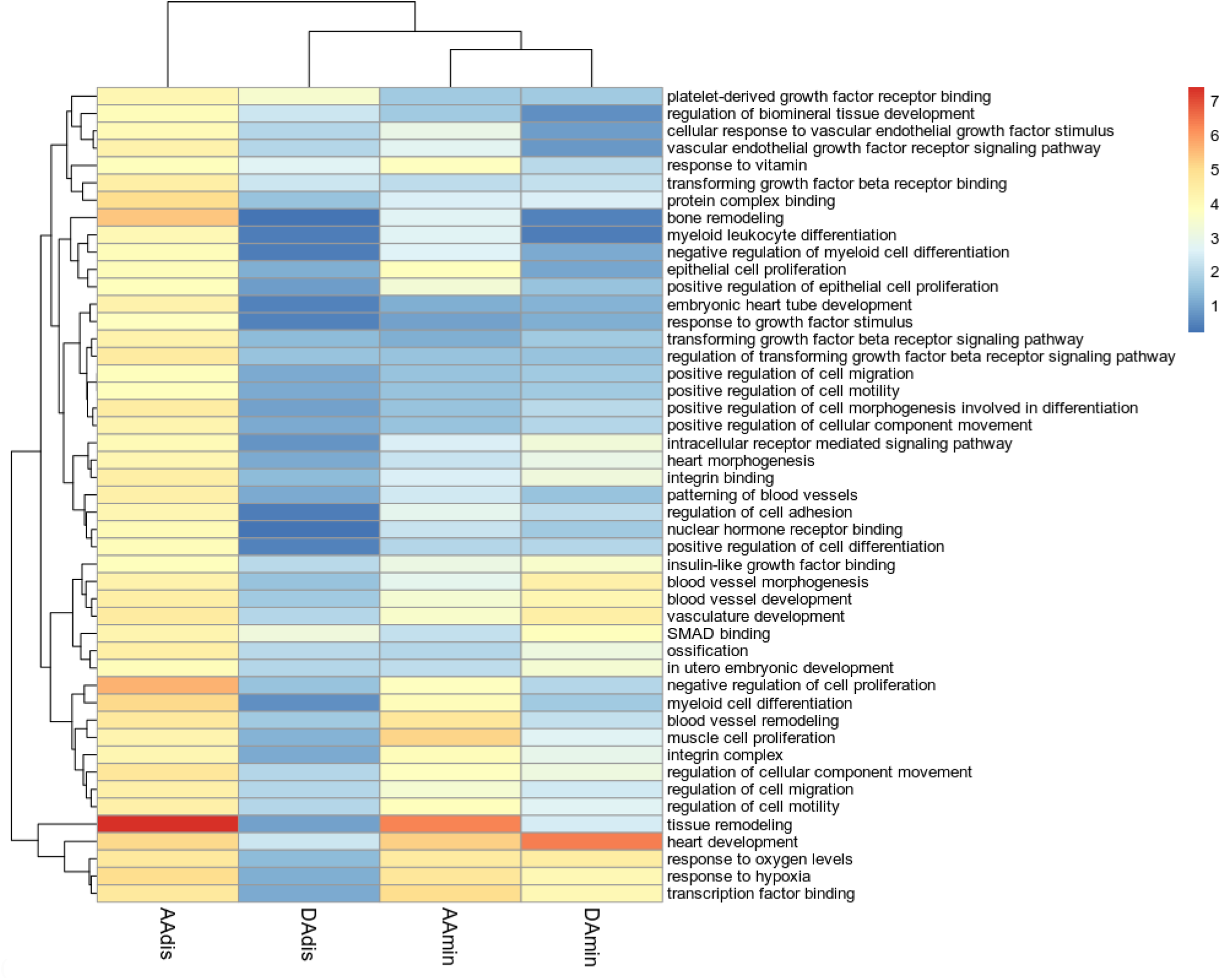
Heatmap of significantly enriched gene ontologies (GO terms) for minimum area and distensibility phenotypes generated by DEPICT. Colour scale denotes the significance of enrichment, -log10 scale. Only GO terms significantly enriched in association with AAdis (FDR<0.01) are presented.

### Co-expression network analyses

Using expression data from single cell transcriptomics of the primate aorta^34^, we generated co-expression modules for aortic endothelial and aortic smooth muscle cells. Using our MAGMA (adj. p-value <0.01) and FUMA gene-based associations (see Methods), we generated functional sub-networks for each aortic trait, highly enriched for our significant genes and identified hub genes for modules expressed in aortic endothelial cells and aortic smooth muscle (see Figure 3 and Supplementary Figures S13-S16). These hub genes include genes involved in smooth muscle cell contraction and differentiation (e.g., *ACTB, MYH10, MYL9, NEXN, ARID5B* and *SVIL*), as well as others associated with TGF-β signalling. In endothelial cells, hub genes identified included *EDN1* and *GATA2*.

**FIGURE 3:**
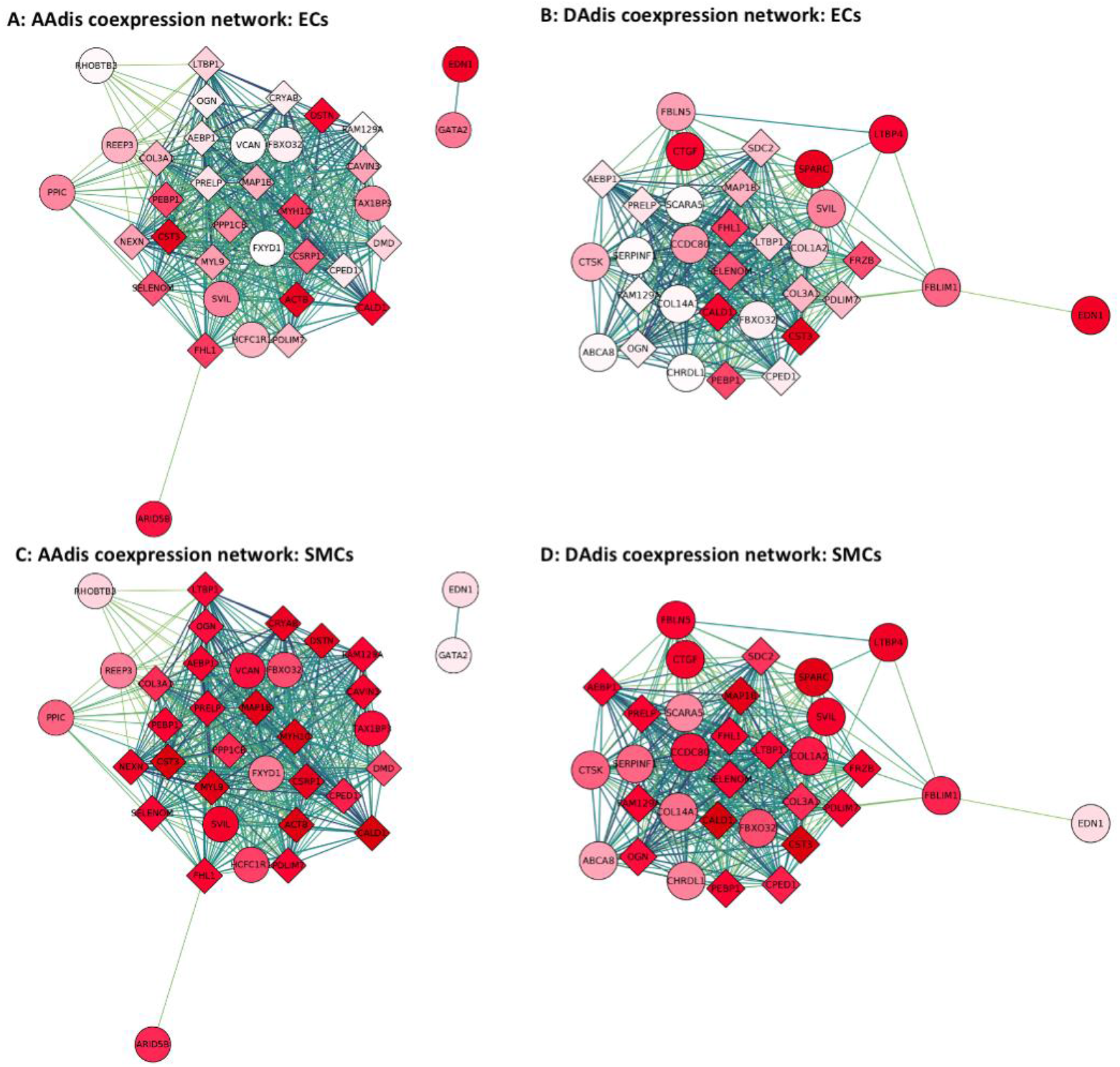
Co-expression networks for aortic distensibility GWAS genes generated with primate single cell expression data for the aorta^34^. The co-expression networks were derived from extended models (r>0.2) in aortic endothelial (AoECs) and aortic smooth muscle cells (AoSMCs). Round circles represent genes which were significantly associated with an aortic trait in the current GWAS. Diamonds represent other genes significantly co-expressed in the published single cell data for the cell-type indicated. The deeper the shade of red, the higher the level of expression of that gene in the specified cell-type. The strength of co-expression is denoted by the colour of the lines joining genes with higher correlations indicated by darker lines. “Hub genes” are found in the centres of these modules. See Supplementary Figures S13-S16 for further co-expression results.

We used these co-expression modules for pathway analyses as described in Methods. The importance of extracellular matrix, vascular smooth muscle cell contraction and developmental pathways were highlighted by enrichment of GO biological pathway and molecular function terms (e.g. extracellular matrix organisation, collagen-containing extracellular matrix, contractile fibre, muscle contraction, actin binding, myosin binding, cardiovascular system development). GO terms related to the TGF-β pathway were also significantly enriched in functional modules derived from gene associations with all the aortic phenotypes (FDR<0.05).

### Phenome-wide association studies and MR-PheWAS

We performed a Phenome Wide Association Study (PheWAS) and a subsequent Mendelian Randomisation – Phenome Wide Association Study (MR-PheWAS) to explore associations between our aortic traits of interest and clinical diagnoses in the whole UK Biobank population for which relevant clinical data were available (n= up to 406,827), controlling for age and sex.

Initial PheWAS identified significant phenotypic associations between aortic traits and multiple hypertension-related clinical codes. Aortic areas showed a positive phenotypic association with hypertension (for example, AAmin β=0.001; p=2.5×10^−21^). Aortic distensibility was negatively associated with hypertension (β=-0.217; p= 1.65×10^−25^). There was a significant negative association for all traits with type II diabetes mellitus.

A subsequent analysis of genotypic associations using MR-PheWAS supported a significant causal relationship between AAdis (MTAG) and aortic aneurysms (Inverse Variance Weighted (IVW) OR=0.28, 95% CI 0.16-0.50; p value 2.14×10^−5^; consistent directions of effect with Weighted Median (WM)/MR Egger and with use of the single trait analysis of AAdis as the genetic instrument, though the latter did not reach significance), suggesting clinical meaningfulness of the distensibility phenotype. MR-PheWAS also suggested that ascending and descending aortic areas are causally related to the risk of aortic aneurysms without evidence of significant pleiotropy.

### Relationship between blood pressure and aortic dimensions

We tested further for bi-directional causal relationships between quantitative blood pressure traits (using GWAS summary statistics from a previous study^35^) and aortic areas using Mendelian randomisation (MR). MR results supported a bidirectional causal relationship between ascending aortic areas and diastolic blood pressure (DBP; AAmin->DBP; β_IVW_=0.004, p=4.3×10^−16^; DBP->AAmin: β_IVW_=6.6; p=2.5×10^−5^) and between ascending aortic area and pulse pressure (PP; AAmin->PP: β_IVW_=-0.007; p=7.1×10^−15^; PP-> AAmin: β_IVW_=-8.1, p=4.4×10^−7^). MR-Egger estimates were consistent for all but the DBP-> AAmin analysis, for which the estimates were in the opposite direction. Contamination mixture MR (MR-ConMix, see Methods) showed consistent findings for all analyses. Similar analyses for causal relationships with blood pressure were not performed for distensibility since blood pressure is used for calculation of the trait.

### Genetic relationships between aortic traits and cerebral small vessel disease or cervical artery dissection

We explored genetic correlations and potential causal relationships between aortic traits and brain SVD estimated from the brain MRI measure of white matter hyperintensity (WMH) burden. Using LDSC, we identified a significant genetic overlap between all aortic traits and WMH burden which defined a positive association with minimum aortic area and an inverse association for the distensibility measures (AAmin r_g_=0.20, p=0.001; DAmin r_g_=0.22, p=2.19×10^−5^; AAdis r_g_=-0.22, p=5.0×10^−4^; DAdis r_g_=-0.33; p=1.0×10^−4^). In further analyses we found no significant genome-wide overlap between aortic traits and risks of cervical artery dissection, a leading cause of stroke in young people which was associated with aortic phenotypes in an earlier study^36^. However, the regional level overlap estimates from a Bayesian pairwise GWAS (GWAS-PW) suggest a high probability of shared variants between aortic distensibility traits and the cervical artery dissection (CeAD) risk locus *PHACTR1-EDN1*. Most of the other CeAD risk loci with high probability of shared variants with aortic traits harbour single nucleotide variants (SNVs) associated at genome-wide significance with one or more of the aortic traits. Exceptions to this include regions at chromosomes 12 (including *c12orf49, RNFT2, PAWR, OTOGL*), 16 (including *CMIP, PKD1L2, BCO1)* and 2 (including *MBD5, EPC2, LYPD6B*), suggesting further novel biologically relevant associations with aortic traits in these regions.

### Exploring relationships between aortic traits and white matter hyperintensity burden using Mendelian randomisation

A lower aortic distensibility or a greater ascending aortic area is genetically correlated with increased burden of WMH in our data. We hypothesised that there might be a causal relationship between these aortic traits and cerebral small vessel disease. Although a two-sample MR using genetic associations with aortic traits as the instrumental variable and WMH as the outcome showed no evidence for a causal association for any of the aortic traits after FDR correction, after accounting for the effect of blood pressure (either systolic or pulse pressure) using a multivariable MR, we found evidence for a direct causal effect of both ascending and descending aortic distensibility and ascending aortic area on WMH burden. Lower distensibility and higher area were associated with an increased WMH burden (for AAdis β=-0.12, p= 1.49×10^−3^ and DAdis β=-0.21, p=1.14 ×10^−3^ using MTAG-derived associations as the instrumental variable and for AAmin β=3.8×10^−4^, p=2.91×10^−3^ using stage 1 associations as the instrumental variable).

## DISCUSSION

Our analysis provides a first large-scale GWAS of ascending and descending aortic distensibilities, and adds substantively to the literature concerning the genetic basis of variation in aortic dimensions. We show that aortic distensibility has a significant heritable component, with 11% of the variance in AA distensibility and 10% of the variance in DA distensibility explained by the common genetic variants included in our study (increasing to 24% and 21% respectively for the MTAG analysis). We identify 38 loci associated with these measures of aortic stiffness, and a total of 31 novel loci for aortic traits including aortic areas (102 loci overall). Annotation of these loci provides evidence for mechanistic associations of TGF-β, IGF, PDGF and VEGF signalling pathways with aortic distensibility.

The clinical significance of our findings is suggested by the potential causal associations between AAdis (and other aortic traits) and aortic aneurysms defined by MR-PheWAS. Multivariable MR provides new evidence for possible mechanistic associations between cerebral small vessel disease and both aortic distensibilities and aortic area, helping to explain their long-recognised clinical relationships^20, 37^.

Mendelian aortopathy or cardiovascular disease associated genes *ELN*^*38*^, *THSD4*^*39*^, *FBLN5*^*40, 41*^, *PRDM6*^*42*^ and *ABCC9*^*43*^ directly overlap loci associated with aortic distensibility phenotypes and thus are strong candidates for expression of functional effects of variation at the corresponding loci. Similarly, Mendelian disease genes *FBN1*^*44*^, *MYH7*^*45*^, *TBX20*^*46*^, *MASP1*^*47*^ *and LOX*^*48*^ overlap loci associated with aortic area phenotypes. Other genes associated with aortic area in our analyses have previously been associated with risks of acute aortic dissection (*ULK4, LRP1*^*49*^). Several gene ontologies were associated with our measured aortic traits which are also of significance in Mendelian aortic disease^50^. Those GO terms related to the extracellular matrix, cardiovascular development and vascular smooth muscle cell function (with genes such as *ELN, ABCC9, ANO1* and *PRDM6* associated with AAdis) were amongst the most consistently identified. This overlapping genetic landscape of distensibility (and aortic areas) and aortic aneurysms, and our finding of a likely causal link between these phenotypes using MR-PheWAS suggest that functional pathways related to genes associated with quantitative aortic traits contribute to the pathogenesis of aortic disease. Together, these observations also support the growing consensus that cardiovascular disease phenotypes may be expressed as a result of extremes of normal genetic variation in the population^59^, and support the use of distensibility to predict both aneurysm formation and progression^4^.

Our data also support a role for TGF-β signalling in determining aortic distensibility in the general population. The TGF-β pathway has long been recognised as an important modulator of aortic function, with variants in many of the major components identified as causal in Mendelian aortic disease such as Loeys-Dietz syndrome^51^. Gene ontologies and cell-specific co-expression modules associated with the measured aortic traits suggest that IGF, PDGF and VEGF signalling also play significant roles in aortic biology. The roles of insulin and of insulin-like growth factor signalling are of considerable therapeutic interest for aortic pathology given recent evidence suggesting that metformin, a known regulator of both signalling pathways^54^, could be an effective treatment for abdominal aortic aneurysm, and the consequent initiation of clinical trials testing this^55, 56^.

Each of these gene sets offers interesting candidates in the search for new Mendelian aortic disease genes. Our results also add to the literature on sex differences in the genomic regulation of cardiovascular traits^57, 58,^ with new evidence presented here suggesting distinct, biologically relevant associations in males and females and implicating genes such as *ADAMTS7, SCAI, ALDH2* and *FES* as sex-specific determinants of aortic traits and thus possibly also the related diseases.

The strongest SNV association with ascending aortic distensibility was found in close proximity and upstream of *ELN*, the gene encoding elastin, a functionally central component of aortic elastic fibres. *FBLN5* was strongly associated with DAdis. This encodes fibulin-5, another key extracellular matrix protein and a mediator of elastic fibre assembly^63^. Variants in *FBLN5* cause a Mendelian form of *cutis laxa* associated with aortic aneurysm, vascular tortuosity and supravalvular aortic stenosis^40, 41^. The importance of extracellular matrix (ECM) composition and regulation is also demonstrated by the identification of multiple ECM-related GO terms associated with aortic phenotypes. Specific gene associations also serve to emphasise this, including three members of the ADAMTS family, which regulate ECM turnover: *ADAMTS7* and *ADAMTS8*, both previously associated with aortic minimum areas and replicated here, and a novel association of ascending aortic distensibility with *ADAMTS9*. The strong association of AAdis with *HAS2* (encoding a hyaluronan synthetase) demonstrated that glycosaminoglycan components of the ECM are also key determinants of aortic traits.

Other specific associations provide insights into the complexity of aortic biology. For example, the second most significant association with AAdis is within a long, non-coding RNA (lncRNA) just 3’ of *PCSK1*, a proprotein convertase whose substrates include many hormones such as renin, insulin and somatostatin (associated previously with body mass index and obesity^64^) and therefore which may mediate multiple endocrine influences on aortic traits. The third most significant locus associated with AAdis was previously associated with coronary artery disease^65^ and spans *SMG6* (a regulator of nonsense mediated decay) and *SRR*, a serine racemase. The causal gene at this locus is thought to be *SMG6*, although functional data demonstrating strong eQTLs for *SRR* in all the risk alleles identified suggests it remains a candidate gene for this locus.

The shared genetic basis of ascending and descending distensibilities is limited (Supplementary Figure 4), consistent with the differing developmental origins of these parts of the aorta, and associated differences in structures of the aortic wall, in which elastin content diminishes more distally^66^. The most significant association for descending aortic distensibility is in *PLCE1*, a gene previously associated with blood pressure traits^35, 67^. Evidence from knockout mice suggests that PLCE1 contributes to the integration of β-adrenergic signalling with inputs from IGF-1 and other pathways to regulate cardiomyocyte differentiation and growth. We speculate that it might play a similar integrative role in the development and remodelling of aortic tissues.

Associations between aortic traits and brain small vessel disease have long been recognised, but the mechanisms responsible have not been defined clearly^20, 68^. This has been a particularly difficult relationship to untangle, as both are subject to confounding influences of blood pressure and other pleotropic factors. Our multivariate MR provides novel evidence suggesting that aortic traits including distensibility are causally linked to WMH and that this relationship is independent of (and additive to) the effects of blood pressure. By inference, as WMH burden predicts cognitive decline and dementia (with evidence supporting a causal association with Alzheimer type dementia^16, 37^), these results indirectly suggest that aortic stiffening could also contribute to cognitive decline and dementia, e.g. through altered haemodynamics and resultant changes in cerebral blood flow leading to effects on brain endothelial cell function and small vessel remodelling^69, 70^.

A shared genomic influence on aortic distensibility and cervical artery dissections was identified at the *PHACTR1/EDN1* locus. Previously, this locus was implicated in coronary artery disease^71^, myocardial infarction^72^, migraine^73^, fibromuscular dysplasia^74^ and cervical artery dissection^75^. We demonstrated an association of *EDN1* with ascending aortic distensibility, and further characterised *EDN1* as a hub gene in co-expression networks derived from aortic endothelia, suggesting that *EDN1* may be responsible for (or functionally contribute to) this shared genetic association with both CeAD risk and aortic distensibility,

Although we have made several novel observations, there are obvious limitations of our study. The GWAS was restricted to the analysis of Caucasian individuals, and so the generalisability of our results for people from other ethnic backgrounds is uncertain. Second, the accuracy (and possibly also the precision) of the distensibility measures likely was reduced by the need to use non-invasive blood pressure measurements (acquired on the same day as the imaging) as proxies for central blood pressure recordings. The ascending and descending aortic distensibility measures also suffer from confounding due to a likely bidirectional relationship with blood pressure, given the use of blood pressure indices in the derivation of the phenotype. Uncontrolled confounding from residual effects of blood pressure could bias the Mendelian randomization analyses. Nevertheless, our genetic associations were significantly enriched for genes expressed in the aorta and identify genes known to be important in aortic biology, affording some confidence in the robustness of our results.

In summary, our results provide genetic association data highlighting roles for TGF-β and other growth factor (IGF, PDGF, VEGF) signaling pathways in the elastic function of the aorta and, by inference, in aortic disease. We present new evidence for potential causal links between lower aortic distensibility and increased risk of aortic aneurysm and for common causal mechanisms relating cerebral small vessel disease and aortic structure and function that could explain the clinically observed relationships between late life cognitive decline and aortic disease^14, 18^. Better understanding of the underlying mechanisms based on these genetic data could lead to the identification of new therapeutic targets for reduction of both cardiovascular disease and dementia risks.

## METHODS

### Data

The UK Biobank cardiac magnetic resonance imaging (CMR) was conducted using a rigorously controlled acquisition protocol^76^. The mean age at the time of CMR was 64 ± 8 years (range 45–82, 49% of participants were male). Exclusion criteria for imaging included a range of relative contraindications to magnetic resonance imaging scanning as well as childhood onset disease and pregnancy. Aortic cine images were acquired using transverse bSSFP sequence at the level of the pulmonary trunk and right pulmonary artery on clinical wide bore 1.5 Tesla scanners (MAGNETOM Aera, Syngo Platform VD13A, Siemens Healthcare). Each cine image sequence consists of 100 time frames. The typical image size is 240×196 pixel with the spatial resolution of 1.6×1.6 mm^2^. Brachial blood pressure was obtained using a manual sphygmomanometer and converted into central blood pressure for the distensibility calculations by applying a brachial-to-aortic transfer function using the Vicorder software^76^.

### Derivation of imaging phenotypes

A recurrent convolutional neural network was developed for aortic image segmentation and trained using manual annotations of 800 ascending aorta and descending aorta images (400 subjects and 2 time frames per subject)^25, 27^ The network was applied to segmenting aortic images across the cardiac cycle. A semi-automated quality control was performed for all segmentations, consisting of automated checking of missing or fragmented segmentation and subsequent manual checking on segmentation screenshots. Six aortic imaging phenotypes were calculated based on the automated segmentations, including those for the maximal area, minimal area and distensibility for both the ascending aorta and descending aorta^4^. Distensibility was calculated as

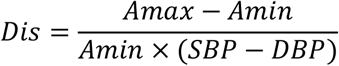

where Amax and Amin denote the maximal and minimal area and SBP and DBP denote the systolic and diastolic central blood pressure, measured at the imaging visit during the study protocol (measured brachially and converted to central blood pressure by applying a brachial-to-central transfer function as described above). After running the image segmentation pipeline and performing quality control, imaging phenotypes were available for 36,996 participants.

### Genomic analyses

We performed stage 1 GWAS on six imaging phenotypes (AAmax, AAmin, DAmax, DAmin, AAdis and DAdis). Outliers with phenotype values >4 standard deviations (SDs) from the mean were excluded to ensure we did not include patients with undiagnosed aortic aneurysm in our results. After exclusions for image quality control, outlying BMI (<15 or >40), stage IV hypertension, missing covariates, diagnosis of aortic disease and non-white ethnicity, 4,406 participants were excluded leaving 32,590 individuals for the GWAS of AAmax, AAmin, DAmax, and DAmin, and 29,895 for the GWAS of AAdis and DAdis (the latter figure is lower due to more missing contemporaneous blood pressure recording data and incomplete imaging sets). See Supplementary Figure S1 for more details on exclusions. After exclusions, we rank-normalised the distensibility phenotypes due to the non-normal distribution of the distensibility phenotypes (Supplementary Figure S2). Aortic area phenotypes approximated a normal distribution and so raw areas were used to facilitate interpretation of the effect sizes. The genetic model was adjusted for age at time of imaging, sex, mean arterial pressure, height, and weight. We used the linear mixed model approach implemented in BOLT-LMM (v2.3.4)^77^. The genetic relationship matrix (GRM) constructed by BOLT was based on all directly genotyped SNPs (N = 340,336) passing the threshold settings (MAF > 0.05, p(HWE) > 1e^-6^ and genotype calling rate > 98.5). For the main analysis, a threshold of MAF >0.01 was applied to the SNPs. Genomic inflation (lambda) was calculated in R as the median chi square values derived from the p-values divided by the expected median of a chi square distribution with 1 degree of freedom.

We used MTAG (version 1.0.8)^28^ for multi-trait analysis of GWAS summary statistics to increase power. MTAG can identify genetic loci associated with a particular phenotype where the single-trait GWAS is underpowered. The method uses the correlation structure of the trait in question with other traits to boost power. It therefore may fail to identify trait-specific loci but will increase power to detect loci associated with the other related and correlated traits. We used all 6 aortic phenotypes for our MTAG analysis. Regression coefficients (beta) and their standard errors were used for MTAG. The results of the multi-trait analyses are shown as Figure 1B and in Supplementary Figures S7-8.

We additionally conducted a sex-specific analysis by performing GWAS on aortic areas for autosomal SNPs in men and women separately, using the BOLT-LMM pipeline as above, and compared the sizes of the sex-specific genetic associations for SNPs with a P-value smaller than 5×10^−8^ using a z-test^78^. We did not repeat this analysis for distensibility phenotypes as it was underpowered because of the reduced cohort size and smaller effect sizes for the distensibility SNPs as well as lower heritability estimates for these traits.

To classify genomic loci associated with our imaging phenotypes, GWAS summary statistics were processed using FUMA (v1.3.6)^32^ and a pre-calculated LD structure based on the European population of the 1000 Genome Project^82^. SNPs that reached genome-wide significance (p<5×10^−8^) and with r^2^<0.6 were defined as independently significant. All variants with r^2^ ≥ 0.6 were labelled as candidate variants for further annotation by FUMA. In a second clumping procedure to define lead SNPS, those correlated with r^2^<0.1 were defined as independently significant. Finally, proximal LD blocks of independent significant SNPs with less than 250kb distance were merged and considered as a single genomic locus. To consolidate genomic loci across different traits, the merge function implemented in bedtools was applied^79^.

For the association of SNPs with genes, biological processes and tissue expression, we applied the SNP2GENE function implemented in FUMA to the summary SNP statistics. For the positional mapping of SNPs to genes, a maximal distance of 10kB was set. eQTL mapping was performed based on the aorta tissue samples in GTEx v8 using only gene pairs with significant SNPs (with the default settings of FDR < 0.05 or p value < 1e^-3^). 3D chromatin interaction mapping was based on HiC aorta data (GSE87112) within a promotor region window of 250bp upstream and 500bp downstream from the transcriptional start site and a threshold for significant loops of FDR < 1e^-6^. Enhancer and promotor regions were annotated using the aorta epigenome (E065) from the Roadmap Epigenome Project (http://www.roadmapepigenomics.org/). MAGMA (Multi-marker Analysis of GenoMic Annotation) was employed to obtain the significance of individual genes with the specificity of tissue expression based on 54 types in GTEx v8 and the association with 10,678 gene sets from MsigDB v6.2 (with 4761 curated gene sets and 5917 GO categories). Further annotation of significant SNPs and loci was performed manually with SNP lookups in GTEx v8^29^; normalised effect sizes are reported from this dataset using “Artery-Aorta” for the main analysis and including other arterial tissue types (“Artery – Tibial” and “Artery – Coronary”) where stated. For Gene Ontology (GO) enrichment analysis, the Data-driven Expression Prioritized Integration for Complex Traits (DEPICT) software was applied (v1 beta rel194, www.broadinstitute.org/depict), which is based on probabilistic memberships of genes across reconstituted gene sets^80^ For LD-based clumping by PLINK^81^, which precedes the DEPICT analysis, a p-value threshold of 10^−5^, a distance threshold of 500kb and a LD threshold of 0.1 was set (following the recommendations on the DEPICT website). Note that DEPICT excludes any SNPs in the HLA region, on a sex chromosome, or not found in the 1000 Genomes Project data.

Comparisons between our data and those reported by Pirruccello et al^30^. and Tcheandjieu et al^31^ were made using the lead SNPs and corresponding beta-values. To assess the degree of convergence of their studies with our results, lead SNPs were assigned to a genomic locus found in our study if they were within the locus or a 250kb distance interval.

### LD Score regression

We performed LD Score regression to assess the heritability of the imaging phenotypes. The genetic correlation between imaging phenotypes and blood pressure by LD score was computed using 1000 Genomes European data^82^. We used the GWAS summary statistics for SBP, DBP and pulse pressure (PP) from the International Consortium for Blood Pressure (ICBP)^35^ for the corresponding analyses for genetic correlation with blood pressure.

### Co-expression analysis

Co-expression networks for aortic phenotypes were derived using a single cell RNA-seq (scRNA-seq) data set for primate arteries^34^. The data were obtained from Gene Expression Omnibus (accession number GSE117715) and included read counts for over 9000 single cells from aortas and coronary arteries of 16 *Macaca fascicularis*. Low abundance genes were removed if they had read counts of less than 5% of the cells, leaving a total of 9903 genes for further analysis. The Bioconductor package *scater* was applied to compute log-transformed normalised expression values from the count matrix^83^. Subsequently, correlation of expression and its significance was derived using the *correlatePairs* function of the Bioconductor *scran* package, which calculated modified Spearman correlation coefficients and derived their significance using a permutation approach^84^. A basal co-expression network was constructed using gene pairs with significant correlation (FDR < 0.01) and a minimum Spearman correlation coefficient *rho* of 0.1 or 0.2. Subsequently, sub-networks for imaging phenotypes were derived by retrieving gene pairs with at least one gene associated by MAGMA or FUMA with the specific phenotype.

Functional enrichment of genes connected with variants identified on GWAS was carried out using overrepresentation enrichment analysis implemented in the Bioconductor *clusterProfiler* package^85^. The background gene set (or universe) was defined by the genes covered by the scRNA-seq data after exclusion of low abundance genes.

### Mendelian randomization (MR)

To investigate potentially causal relationships between aortic phenotypes and diseases, we performed bidirectional two-sample Mendelian randomization (MR) using our GWAS for aortic imaging phenotypes and the International Consortium for Blood Pressure GWAS on blood pressure^35^. We limited our MR analyses to AAmax, AAmin, DAmax, and DAmin because blood pressure is included in the calculation of the distensibility measure.

For either direction of potential causal relationships between the aortic phenotypes and blood pressure, we selected SNPs associated with the exposure at genome-wide significance level (P-value<5×10^−8^ and F-statistic>10). For SNPs correlated with r^2^ greater than 0.1, we used only the SNP with the smallest p-value for the SNP-exposure association. We tested the validity of the genetic variants as instrumental variables using the contamination mixture method (MR-ConMix)^86^. The contamination mixture method constructs a likelihood function based on the SNP-specific estimates and evaluates the SNP-specific contribution to the likelihood.

In each case, we estimated SNP-specific associations as the ratio of SNP outcome to exposure associations (Wald ratio)^87^. SNP-specific associations were combined using the inverse-variance weighted (IVW) estimator ^88^. The putative causal effect (β_IVW_) of an exposure on a given outcome was estimated using the inverse-variance weighting (IVW) method as the weighted sum of the ratios of beta-coefficients from the SNP-outcome associations for each variant (j) over corresponding beta-coefficients from the SNP-exposure associations (β_j_). The ratio estimates from each genetic variant were averaged after weighting on the inverse variance (W_j_) of β_j_ across L uncorrelated SNPs,

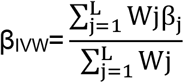

We also used weighted median (WM) and MR-Egger regressions as sensitivity methods to test the robustness of associations ^88^. Potential horizontal pleiotropic effects were investigated using MR-Egger ^89^. Outlier SNPs identified by MR-PRESSO were excluded from the analyses ^90^. In an additional analysis, we tested the validity of the genetic variants as instrumental variables using MR-ConMix^86^. We accounted for multiple comparisons of four aortic imaging phenotypes, three blood pressure traits, and two directions using Bonferroni correction with a P-value threshold of 0.05/(4*3*2)=0.002

MR analysis was also used to investigate the potential causal relationships between different aortic traits with WMH. In addition to IVW, WM, and MR-Egger, we implemented R package RadialMR (available through CRAN repositories)^91^. A p-value < 0.01 correcting for 6 tests (for the 6 aortic traits) was considered significant. In the presence of heterogeneity (P_Het_<0.01, Cochran’s Q statistic) due to horizontal pleiotropy, RadialMR was used in the identification of pleiotropic SNPs that have the largest contribution to the global Cochran’s Q statistic by regressing the predicted causal estimate against the inverse variance weights. After excluding influential outlier SNPs, the IVW test was repeated along with MR-Egger regression in which the regression model contains the intercept term representing any residual pleiotropic effect ^92^. Non-significant MR-Egger intercept was used as an indicator to formally rule out horizontal pleiotropy. Relative goodness of fit of the MR-Egger effect estimates over the IVW approach was quantified using Q_R_ statistics, which is the ratio of the statistical heterogeneity around the MR-Egger fitted slope divided by the statistical heterogeneity around the IVW slope. A Q_R_ close to 1 indicates that MR-Egger is not a better fit to the data and therefore offers no benefit over IVW ^91^.

### Multivariable MR

We also conducted multivariable two sample MR (MVMR) to estimate the direct effect of aortic traits on the cerebral small vessel disease (cSVD) outcome (WMH) after accounting for potential confounding with blood pressure traits, by conditioning on every other explanatory variable included in the model. Different combinations of explanatory variables were considered and the F_TS_ conditional on the other variables was calculated as a measure of instrument strength ^93^. Briefly, MVMR by regressing a given instrumental variable on all the remaining variables as controls generates a predicted value for the instrumental variable that is not correlated with other variables in the model thus accounting for possible pleiotropic effects.

### MR phenome-wide association studies (MR-PheWAS)

We performed an MR-PheWAS using data from UK Biobank participants who did not undergo aortic imaging to assess the effects of aortic traits on clinical disease classifications. Using the PheWAS package (https://github.com/PheWAS/PheWAS), we mapped 1,157 phecodes with more than 200 cases from the International Classification of Diseases, 9th Revision, Clinical Modification (ICD-9-CM) and the International Classification of Diseases, 10th Revision, Clinical Modification (ICD-10-CM)^.94^ For each aortic imaging phenotype, we selected SNPs with a p-value<5×10^−8^ and minor allele frequency > 0.05 from the single trait GWAS for AAmax, AAmin, DAmax, and DAmin, and from the multi trait GWAS for AAdis and DAdis. Given that we considered a large number of phecodes as the outcome in the MR-PheWAS analysis with the number of cases ranging from 201 to 116,879 (median=1,353), we used a more stringent LD threshold for independent SNPs (r^2^<0.01) to achieve a better stablised model. The PheWAS model was adjusted for age, sex, genotype array, and its four principal components for population stratification. MR estimates were then obtained for each pair of aortic traits and phecode by combining the SNP-specific associations using IVW, WM, and MR-Egger. We accounted for multiple comparisons of 1,157 phecodes using Bonferroni correction with p-value<0.05/1,157 =4.3×10^−5^.

### Analyses of associations of aortic and cervical artery dissection

LD score regression (LDSR) method was applied to test genetic correlation at the genome-wide scale for the different aortic traits with the most common MRI feature of cSVD, WMH and with cervical artery dissection (CeAD). GWAS summary statistics were obtained from recently published consortia GWAS of cerebral phenotypes (from the CHARGE and CADISP consortia respectively)^37,75^. For this, common variants, mapping to the Hapmap3 reference panel were employed. As the slope from the regression of the Z-score product from the two GWAS summary statistics on the LD-score gives the genetic covariance, the intercept of the genetic covariance was used as an indirect measure of sample overlap ^95^, which corresponds to the average polygenic effects captured by genetic variants spread across the genome. A p-value < 0.006 (adjusting for 8 simultaneous tests) was considered significant.

LDSR could potentially miss significant correlations at the regional level due to the balancing-effect^96^. A Bayesian pairwise GWAS approach (GWAS-PW) was applied to systematically test for locally correlated regions^97^. The GWAS-PW identified trait pairs with high posterior probability of association (PPA) using a shared genetic variant (model 3, PPA3> 0.90). To ensure that PPA3 is unbiased by sample overlap, fgwas v.0.3.6^98^ was run on each pair of traits and the correlation estimated from regions with null association evidence (PPA3<0.20) was used as a correction factor. Additionally, to estimate the directionality of associations between trait pairs in regions with PPA3>0.90, a simple rank-based correlation test was applied. Independence between regions was estimated as proposed by Berisa and Pickrell^99^. Only the most strongly associated variant for the outcome per region showing high PPA3 is reported.

## Data Availability

Summary statistics for all GWAS analyses are available upon request.

## SUPPLEMENTARY FIGURES

Supplementary Figure 1 Exclusions

Supplementary Figure 2 Distribution of traits

Supplementary Figure 3 Phenotypic correlation between traits

Supplementary Figure 4 Genotypic correlation between traits (LDSC)

Supplementary Figure 5 All Manhattan plots stage 1 GWAS

Supplementary Figure 6 All QQ plots stage 1 GWAS

Supplementary Figure 7 All Manhattan plots MTAG

Supplementary Figure 8 All QQ plots stage 2 (MTAG) GWAS

Supplementary Figure 9 FUMA genes results

Supplementary Figures 10 MAGMA genes Manhattan plots

Supplementary Figure 11 MAGMA tissue expression

Supplementary Figure 12 MAGMA GO heatmap

Supplementary Figure 13 Coexpression networks for AAdis

Supplementary Figure 14 Coexpression networks for DAdis

Supplementary Figure 15 Coexpression networks for AAmin

Supplementary Figure 16 Coexpression networks for DAmin

## ACKNOWLEDGEMENTS

PMM acknowledges generous personal and research support from the Edmond J Safra Foundation and Lily Safra, a National Institute for Health Research (NIHR) Senior Investigator Award, the UK Dementia Research Institute and the NIHR Biomedical Research Centre at Imperial College London. CF acknowledges generous support from the BHF Imperial Centre of Research Excellence RE/18/4/34215. This research has been conducted using the UK Biobank Resource under Application Number 18545. PE is director of the Medical Research Council (MRC) Centre for Environment and Health and acknowledges support from the MRC (MR/L01341X/1; MR/ S019669/1). PE is director of the Health Protection Research Unit in Chemical and Radiation Threats and Hazards, funded by the NIHR. PE also acknowledges support from the NIHR Imperial Biomedical Research Centre, the Imperial College British Heart Foundation Centre for Research Excellence (RE/18/4/34215), the UK Dementia Research Institute at Imperial College London (MC_PC_17114), and Health Data Research UK for London. SD is supported by a grant overseen by the French National Research Agency (ANR) as part of the ‘Investment for the Future’ Programme ANR-18-RHUS-002 and by the ERC and the EU H2020 under grant agreements 640643, 667375, and 754517.

The CADISP study has been supported by Inserm, Lille 2 University, Institut Pasteur de Lille and Lille University Hospital and received funding from the ERDF (FEDER funds) and Région Nord-Pas de Calais in the frame of Contrat de Projets Etat-Region 2007-2013 Région Nord-Pas-de-Calais - Grant N°09120030, Centre National de Genotypage, Emil Aaltonen Foundation, Paavo Ilmari Ahvenainen Foundation, Helsinki University Central Hospital Research Fund, Helsinki University Medical Foundation, Päivikki and Sakari Sohlberg Foundation, Aarne Koskelo Foundation, Maire Taponen Foundation, Aarne and Aili Turunen Foundation, Lilly Foundation, Alfred Kordelin Foundation, Finnish Medical Foundation, Orion Farmos Research Foundation, Maud Kuistila Foundation, the Finnish Brain Foundation, Biomedicum Helsinki Foundation, Projet Hospitalier de Recherche Clinique Régional, Fondation de France, Génopôle de Lille, Adrinord, Basel Stroke-Funds, Käthe-Zingg-Schwichtenberg-Fonds of the Swiss Academy of Medical Sciences, Swiss Heart Foundation.

## CADISP list of investigators

Belgium: Departments of Neurology, Erasmus University Hospital, Brussels and Laboratory of Experimental Neurology, ULB, Brussels (Shérine Abboud, Massimo Pandolfo); Department of Neurology, Leuven University Hospial (Vincent Thijs). France: Departments of Neurology, Lille University Hospital-Inserm U1171 (Didier Leys, Marie Bodenant), Sainte-Anne University Hospital, Paris (Fabien Louillet, Emmanuel Touzé, Jean-Louis Mas), Pitié-Salpêtrière University Hospital, Paris (Yves Samson, Sara Leder, Anne Léger, Sandrine Deltour, Sophie Crozier, Isabelle Méresse), Amiens University Hospital (Sandrine Canaple, Olivier Godefroy), Dijon University Hospital (Maurice Giroud, Yannick Béjot), Besançon University Hospital (Pierre Decavel, Elizabeth Medeiros, Paola Montiel, Thierry Moulin, Fabrice Vuillier); Inserm U744, Pasteur Institute, Lille (Jean Dallongeville). Finland : Department of Neurology, Helsinki University Central Hospital, Helsinki (Antti J Metso, Tiina Metso, Turgut Tatlisumak); Germany: Departments of Neurology, Heidelberg University Hospital (Caspar Grond-Ginsbach, Christoph Lichy, Manja Kloss, Inge Werner, Marie-Luise Arnold), University Hospital of Ludwigshafen (Michael Dos Santos, Armin Grau); University Hospital of München (Martin Dichgans); Department of Rehabilitation: Schmieder-Klinik, Heidelberg (Constanze Thomas-Feles, Ralf Weber, Tobias Brandt). Italy: Departments of Neurology: Brescia University Hospital (Alessandro Pezzini, Valeria De Giuli, Filomena Caria, Loris Poli, Alessandro Padovani), Milan University Hospital (Anna Bersano, Silvia Lanfranconi), University of Milano Bicocca, San Gerardo Hospital, Monza, Italy (Simone Beretta, Carlo Ferrarese), Milan Scientific Institute San Raffaele University Hospital (Giacomo Giacolone); Department of Rehabilitation, Santa Lucia Hospital, Rome (Stefano Paolucci). Switzerland: Department of Neurology, Basel University Hospital (Philippe Lyrer, Stefan Engelter, Felix Fluri, Florian Hatz, Dominique Gisler, Leo Bonati, Henrik Gensicke, Margareth Amort). UK: Clinical Neuroscience, St George’s University of London (Hugh Markus). USA : Department of Neurology, Salt Lake City, USA (Jennifer Majersik); Department of Neurology, University of Virginia, Charlottesville, USA (Bradford Worrall, Andrew Southerland); Department of Neurology, Baltimore, USA (John Cole, Steven Kittner)

## CO-AUTHOR CONTRIBUTIONS

CF, MF, JH, SF, AD and PMM co-designed the study. CF, MF, WB, JH, AP, AR and MS performed quality control and core analyses. SD, EP, JW and PE provided additional methodological guidance. CF, MF, JH, AD and PMM drafted the manuscript. All authors reviewed, contributed to serial revisions and approved the manuscript.

## References

1. Ohyama Y, Redheuil A, Kachenoura N, Ambale Venkatesh B, Lima JAC. Imaging Insights on the Aorta in Aging. Circulation Cardiovascular imaging. 2018;11(4):e005617.

2. Teixido-Tura G, Redheuil A, Rodriguez-Palomares J, Gutierrez L, Sanchez V, Forteza A, et al. Aortic biomechanics by magnetic resonance: early markers of aortic disease in Marfan syndrome regardless of aortic dilatation? International journal of cardiology. 2014;171(1):56–61.

3. de Wit A, Vis K, Jeremy RW. Aortic stiffness in heritable aortopathies: relationship to aneurysm growth rate. Heart, lung & circulation. 2013;22(1):3–11.

4. Nollen GJ, Groenink M, Tijssen JG, Van Der Wall EE, Mulder BJ. Aortic stiffness and diameter predict progressive aortic dilatation in patients with Marfan syndrome. European heart journal. 2004;25(13):1146–52.

5. Redheuil A, Wu CO, Kachenoura N, Ohyama Y, Yan RT, Bertoni AG, et al. Proximal Aortic Distensibility Is an Independent Predictor of All-Cause Mortality and Incident CV Events: The MESA Study. Journal of the American College of Cardiology. 2014;64(24):2619–29.

6. Mattace-Raso FU, van der Cammen TJ, Hofman A, van Popele NM, Bos ML, Schalekamp MA, et al. Arterial stiffness and risk of coronary heart disease and stroke: the Rotterdam Study. Circulation. 2006;113(5):657–63.

7. Ben-Shlomo Y, Spears M, Boustred C, May M, Anderson SG, Benjamin EJ, et al. Aortic pulse wave velocity improves cardiovascular event prediction: an individual participant meta-analysis of prospective observational data from 17,635 subjects. Journal of the American College of Cardiology. 2014;63(7):636–46.

8. Cuspidi C, Facchetti R, Bombelli M, Re A, Cairoa M, Sala C, et al. Aortic root diameter and risk of cardiovascular events in a general population: data from the PAMELA study. Journal of hypertension. 2014;32(9):1879–87.

9. Kamimura D, Suzuki T, Musani SK, Hall ME, Samdarshi TE, Correa A, et al. Increased Proximal Aortic Diameter is Associated With Risk of Cardiovascular Events and All-Cause Mortality in Blacks The Jackson Heart Study. Journal of the American Heart Association. 2017;6(6).

10. Lam CS, Gona P, Larson MG, Aragam J, Lee DS, Mitchell GF, et al. Aortic root remodeling and risk of heart failure in the Framingham Heart study. JACC Heart failure. 2013;1(1):79–83.

11. de Roos A, van der Grond J, Mitchell G, Westenberg J. Magnetic Resonance Imaging of Cardiovascular Function and the Brain: Is Dementia a Cardiovascular-Driven Disease? Circulation. 2017;135(22):2178–95.

12. van Sloten TT, Protogerou AD, Henry RM, Schram MT, Launer LJ, Stehouwer CD. Association between arterial stiffness, cerebral small vessel disease and cognitive impairment: A systematic review and meta-analysis. Neurosci Biobehav Rev. 2015;53:121–30.

13. Qiu C, Winblad B, Viitanen M, Fratiglioni L. Pulse pressure and risk of Alzheimer disease in persons aged 75 years and older: a community-based, longitudinal study. Stroke; a journal of cerebral circulation. 2003;34(3):594–9.

14. Raisi-Estabragh Z, M’Charrak A, McCracken C, Biasiolli L, Ardissino M, Curtis EM, et al. Associations of cognitive performance with cardiovascular magnetic resonance phenotypes in the UK Biobank. European heart journal cardiovascular Imaging. 2021.

15. Moroni F, Ammirati E, Rocca MA, Filippi M, Magnoni M, Camici PG. Cardiovascular disease and brain health: Focus on white matter hyperintensities. Int J Cardiol Heart Vasc. 2018;19:63–9.

16. Debette S, Schilling S, Duperron MG, Larsson SC, Markus HS. Clinical Significance of Magnetic Resonance Imaging Markers of Vascular Brain Injury: A Systematic Review and Meta-analysis. JAMA Neurol. 2019;76(1):81–94.

17. Wardlaw JM, Smith EE, Biessels GJ, Cordonnier C, Fazekas F, Frayne R, et al. Neuroimaging standards for research into small vessel disease and its contribution to ageing and neurodegeneration. Lancet Neurol. 2013;12(8):822–38.

18. Debette S, Markus HS. The clinical importance of white matter hyperintensities on brain magnetic resonance imaging: systematic review and meta-analysis. BMJ. 2010;341:c3666.

19. King KS, Chen KX, Hulsey KM, McColl RW, Weiner MF, Nakonezny PA, et al. White matter hyperintensities: use of aortic arch pulse wave velocity to predict volume independent of other cardiovascular risk factors. Radiology. 2013;267(3):709–17.

20. Mitchell GF, van Buchem MA, Sigurdsson S, Gotal JD, Jonsdottir MK, Kjartansson O, et al. Arterial stiffness, pressure and flow pulsatility and brain structure and function: the Age, Gene/Environment Susceptibility--Reykjavik study. Brain. 2011;134(Pt 11):3398–407.

21. Henskens LH, Kroon AA, van Oostenbrugge RJ, Gronenschild EH, Fuss-Lejeune MM, Hofman PA, et al. Increased aortic pulse wave velocity is associated with silent cerebral small-vessel disease in hypertensive patients. Hypertension. 2008;52(6):1120–6.

22. Cocciolone AJ, Hawes JZ, Staiculescu MC, Johnson EO, Murshed M, Wagenseil JE. Elastin, arterial mechanics, and cardiovascular disease. American journal of physiology Heart and circulatory physiology. 2018;315(2):H189–H205.

23. Duca L, Blaise S, Romier B, Laffargue M, Gayral S, El Btaouri H, et al. Matrix ageing and vascular impacts: focus on elastin fragmentation. Cardiovascular research. 2016;110(3):298–308.

24. Ferrucci L, Fabbri E. Inflammageing: chronic inflammation in ageing, cardiovascular disease, and frailty. Nature reviews Cardiology. 2018;15(9):505–22.

25. Bai W SH, Qin C, Tarroni G, Oktay O, Matthews PM, Rueckert D. Recurrent Neural Networks for Aortic Image Sequence Segmentation with Sparse Annotations. In: Frangi A, Schnabel J, Davatzikos C, Alberola-López C, Fichtinger G (eds) Medical Image Computing and Computer Assisted Intervention – MICCAI 2018. 2018;MICCAI 2018. Lecture Notes in Computer Science, vol 11073. Springer, Cham..

26. Littlejohns TJ, Holliday J, Gibson LM, Garratt S, Oesingmann N, Alfaro-Almagro F, et al. The UK Biobank imaging enhancement of 100,000 participants: rationale, data collection, management and future directions. Nat Commun. 2020;11(1):2624.

27. Bai W, Suzuki H, Huang J, Francis C, Wang S, Tarroni G, et al. A population-based phenome-wide association study of cardiac and aortic structure and function. Nature medicine. 2020;26(10):1654–62.

28. Turley P, Walters RK, Maghzian O, Okbay A, Lee JJ, Fontana MA, et al. Multi-trait analysis of genome-wide association summary statistics using MTAG. Nature genetics. 2018;50(2):229–37.

29. Consortium G. GTEx v8 2021: https://gtexportal.org/home/.

30. Pirruccello J. Deep learning enables genetic analysis of the human thoracic aorta. BioRXiV. 2020;doi https://doi.org/10.1101/2020.05.12.091934.

31. Tcheandjieu C. High heritability of ascending aortic diameter and multi-ethnic prediction of thoracic aortic disease. MedRXiV. 2021;doi: https://doi.org/10.1101/2020.05.29.20102335.

32. Watanabe K, Taskesen E, van Bochoven A, Posthuma D. Functional mapping and annotation of genetic associations with FUMA. Nature communications. 2017;8(1):1826.

33. de Leeuw CA, Mooij JM, Heskes T, Posthuma D. MAGMA: generalized gene-set analysis of GWAS data. PLoS computational biology. 2015;11(4):e1004219.

34. Zhang W, Zhang S, Yan P, Ren J, Song M, Li J, et al. A single-cell transcriptomic landscape of primate arterial aging. Nature communications. 2020;11(1):2202.

35. Evangelou E, Warren HR, Mosen-Ansorena D, Mifsud B, Pazoki R, Gao H, et al. Genetic analysis of over 1 million people identifies 535 new loci associated with blood pressure traits. Nature genetics. 2018;50(10):1412–25.

36. Tzourio C, Cohen A, Lamisse N, Biousse V, Bousser MG. Aortic root dilatation in patients with spontaneous cervical artery dissection. Circulation. 1997;95(10):2351–3.

37. Sargurupremraj M, Suzuki H, Jian X, Sarnowski C, Evans TE, Bis JC, et al. Cerebral small vessel disease genomics and its implications across the lifespan. Nature communications. 2020;11(1):6285.

38. Kozel BA, Barak B, Kim CA, Mervis CB, Osborne LR, Porter M, et al. Williams syndrome. Nat Rev Dis Primers. 2021;7(1):42.

39. Elbitar S, Renard M, Arnaud P, Hanna N, Jacob MP, Guo DC, et al. Pathogenic variants in THSD4, encoding the ADAMTS-like 6 protein, predispose to inherited thoracic aortic aneurysm. Genetics in medicine : official journal of the American College of Medical Genetics. 2021;23(1):111–22.

40. Van Maldergem L, Loeys B. FBLN5-Related Cutis Laxa. In: Pagon RA, Adam MP, Ardinger HH, Bird TD, Dolan CR, Fong CT, et al., editors. GeneReviews(R). Seattle (WA)1993.

41. Loeys B, Van Maldergem L, Mortier G, Coucke P, Gerniers S, Naeyaert JM, et al. Homozygosity for a missense mutation in fibulin-5 (FBLN5) results in a severe form of cutis laxa. Human molecular genetics. 2002;11(18):2113–8.

42. Li N, Subrahmanyan L, Smith E, Yu X, Zaidi S, Choi M, et al. Mutations in the Histone Modifier PRDM6 Are Associated with Isolated Nonsyndromic Patent Ductus Arteriosus. American journal of human genetics. 2016;99(4):1000.

43. Hiraki Y, Miyatake S, Hayashidani M, Nishimura Y, Matsuura H, Kamada M, et al. Aortic aneurysm and craniosynostosis in a family with Cantu syndrome. American journal of medical genetics Part A. 2014;164A(1):231–6.

44. Dietz HC, Cutting GR, Pyeritz RE, Maslen CL, Sakai LY, Corson GM, et al. Marfan syndrome caused by a recurrent de novo missense mutation in the fibrillin gene. Nature. 1991;352(6333):337–9.

45. Walsh R, Rutland C, Thomas R, Loughna S. Cardiomyopathy: a systematic review of disease-causing mutations in myosin heavy chain 7 and their phenotypic manifestations. Cardiology. 2010;115(1):49–60.

46. Kirk EP, Sunde M, Costa MW, Rankin SA, Wolstein O, Castro ML, et al. Mutations in cardiac T-box factor gene TBX20 are associated with diverse cardiac pathologies, including defects of septation and valvulogenesis and cardiomyopathy. American journal of human genetics. 2007;81(2):280–91.

47. Rooryck C, Diaz-Font A, Osborn DP, Chabchoub E, Hernandez-Hernandez V, Shamseldin H, et al. Mutations in lectin complement pathway genes COLEC11 and MASP1 cause 3MC syndrome. Nature genetics. 2011;43(3):197–203.

48. Guo DC, Regalado ES, Gong L, Duan X, Santos-Cortez RL, Arnaud P, et al. LOX Mutations Predispose to Thoracic Aortic Aneurysms and Dissections. Circulation research. 2016;118(6):928–34.

49. Guo DC, Grove ML, Prakash SK, Eriksson P, Hostetler EM, LeMaire SA, et al. Genetic Variants in LRP1 and ULK4 Are Associated with Acute Aortic Dissections. American journal of human genetics. 2016;99(3):762–9.

50. Pyeritz RE. Heritable thoracic aortic disorders. Current opinion in cardiology. 2014;29(1):97–102.

51. MacCarrick G, Black JH, 3rd, Bowdin S, El-Hamamsy I, Frischmeyer-Guerrerio PA, Guerrerio AL, et al. Loeys-Dietz syndrome: a primer for diagnosis and management. Genetics in medicine: official journal of the American College of Medical Genetics. 2014;16(8):576–87.

52. Duan C, Xu Q. Roles of insulin-like growth factor (IGF) binding proteins in regulating IGF actions. Gen Comp Endocrinol. 2005;142(1-2):44–52.

53. von der Thusen JH, Borensztajn KS, Moimas S, van Heiningen S, Teeling P, van Berkel TJ, et al. IGF-1 has plaque-stabilizing effects in atherosclerosis by altering vascular smooth muscle cell phenotype. The American journal of pathology. 2011;178(2):924–34.

54. Lei Y, Yi Y, Liu Y, Liu X, Keller ET, Qian CN, et al. Metformin targets multiple signaling pathways in cancer. Chin J Cancer. 2017;36(1):17.

55. Fujimura N, Xiong J, Kettler EB, Xuan H, Glover KJ, Mell MW, et al. Metformin treatment status and abdominal aortic aneurysm disease progression. Journal of vascular surgery. 2016;64(1):46–54 e8.

56. Lareyre F, Raffort J. Metformin to Limit Abdominal Aortic Aneurysm Expansion: Time for Clinical Trials. European journal of vascular and endovascular surgery: the official journal of the European Society for Vascular Surgery. 2021;61(6):1030.

57. Lopes-Ramos CM, Chen CY, Kuijjer ML, Paulson JN, Sonawane AR, Fagny M, et al. Sex Differences in Gene Expression and Regulatory Networks across 29 Human Tissues. Cell Rep. 2020;31(12):107795.

58. Nethononda RM, Lewandowski AJ, Stewart R, Kylinterias I, Whitworth P, Francis J, et al. Gender specific patterns of age-related decline in aortic stiffness: a cardiovascular magnetic resonance study including normal ranges. Journal of cardiovascular magnetic resonance: official journal of the Society for Cardiovascular Magnetic Resonance. 2015;17:20.

59. Tadros R, Francis C, Xu X, Vermeer AMC, Harper AR, Huurman R, et al. Shared genetic pathways contribute to risk of hypertrophic and dilated cardiomyopathies with opposite directions of effect. Nature genetics. 2021;53(2):128–34.

60. Davis EC. Elastic lamina growth in the developing mouse aorta. The journal of histochemistry and cytochemistry: official journal of the Histochemistry Society. 1995;43(11):1115–23.

61. Wahart A, Hocine T, Albrecht C, Henry A, Sarazin T, Martiny L, et al. Role of elastin peptides and elastin receptor complex in metabolic and cardiovascular diseases. The FEBS journal. 2019.

62. Urban Z, Riazi S, Seidl TL, Katahira J, Smoot LB, Chitayat D, et al. Connection between elastin haploinsufficiency and increased cell proliferation in patients with supravalvular aortic stenosis and Williams-Beuren syndrome. American journal of human genetics. 2002;71(1):30–44.

63. Papke CL, Yanagisawa H. Fibulin-4 and fibulin-5 in elastogenesis and beyond: Insights from mouse and human studies. Matrix Biol. 2014;37:142–9.

64. Nead KT, Li A, Wehner MR, Neupane B, Gustafsson S, Butterworth A, et al. Contribution of common non-synonymous variants in PCSK1 to body mass index variation and risk of obesity: a systematic review and meta-analysis with evidence from up to 331 175 individuals. Human molecular genetics. 2015;24(12):3582–94.

65. Consortium CAD, Deloukas P, Kanoni S, Willenborg C, Farrall M, Assimes TL, et al. Large-scale association analysis identifies new risk loci for coronary artery disease. Nature genetics. 2013;45(1):25–33.

66. Baxter BT, Davis VA, Minion DJ, Wang YP, Lynch TG, McManus BM. Abdominal aortic aneurysms are associated with altered matrix proteins of the nonaneurysmal aortic segments. Journal of vascular surgery. 1994;19(5):797-802; discussion 3.

67. International Consortium for Blood Pressure Genome-Wide Association S, Ehret GB, Munroe PB, Rice KM, Bochud M, Johnson AD, et al. Genetic variants in novel pathways influence blood pressure and cardiovascular disease risk. Nature. 2011;478(7367):103–9.

68. O’Rourke MF, Safar ME. Relationship between aortic stiffening and microvascular disease in brain and kidney: cause and logic of therapy. Hypertension. 2005;46(1):200–4.

69. Lacolley P, Regnault V, Segers P, Laurent S. Vascular Smooth Muscle Cells and Arterial Stiffening: Relevance in Development, Aging, and Disease. Physiol Rev. 2017;97(4):1555–617.

70. Jefferson AL, Cambronero FE, Liu D, Moore EE, Neal JE, Terry JG, et al. Higher Aortic Stiffness Is Related to Lower Cerebral Blood Flow and Preserved Cerebrovascular Reactivity in Older Adults. Circulation. 2018;138(18):1951–62.

71. Coronary Artery Disease Genetics C. A genome-wide association study in Europeans and South Asians identifies five new loci for coronary artery disease. Nature genetics. 2011;43(4):339–44.

72. Beaudoin M, Gupta RM, Won HH, Lo KS, Do R, Henderson CA, et al. Myocardial Infarction-Associated SNP at 6p24 Interferes With MEF2 Binding and Associates With PHACTR1 Expression Levels in Human Coronary Arteries. Arteriosclerosis, thrombosis, and vascular biology. 2015;35(6):1472–9.

73. Anttila V, Winsvold BS, Gormley P, Kurth T, Bettella F, McMahon G, et al. Genome-wide meta-analysis identifies new susceptibility loci for migraine. Nature genetics. 2013;45(8):912–7.

74. Kiando SR, Tucker NR, Castro-Vega LJ, Katz A, D’Escamard V, Treard C, et al. PHACTR1 Is a Genetic Susceptibility Locus for Fibromuscular Dysplasia Supporting Its Complex Genetic Pattern of Inheritance. PLoS genetics. 2016;12(10):e1006367.

75. Debette S, Kamatani Y, Metso TM, Kloss M, Chauhan G, Engelter ST, et al. Common variation in PHACTR1 is associated with susceptibility to cervical artery dissection. Nature genetics. 2015;47(1):78–83.

76. Petersen SE, Matthews PM, Francis JM, Robson MD, Zemrak F, Boubertakh R, et al. UK Biobank’s cardiovascular magnetic resonance protocol. Journal of cardiovascular magnetic resonance: official journal of the Society for Cardiovascular Magnetic Resonance. 2016;18:8.

77. Loh PR, Tucker G, Bulik-Sullivan BK, Vilhjalmsson BJ, Finucane HK, Salem RM, et al. Efficient Bayesian mixed-model analysis increases association power in large cohorts. Nature genetics. 2015;47(3):284–90.

78. Altman DG, Bland JM. Interaction revisited: the difference between two estimates. BMJ. 2003;326(7382):219.

79. Quinlan AR, Hall IM. BEDTools: a flexible suite of utilities for comparing genomic features. Bioinformatics. 2010;26(6):841–2.

80. Pers TH, Karjalainen JM, Chan Y, Westra HJ, Wood AR, Yang J, et al. Biological interpretation of genome-wide association studies using predicted gene functions. Nature communications. 2015;6:5890.

81. Purcell S, Neale B, Todd-Brown K, Thomas L, Ferreira MA, Bender D, et al. PLINK: a tool set for whole-genome association and population-based linkage analyses. American journal of human genetics. 2007;81(3):559–75.

82. Bulik-Sullivan BK, Loh PR, Finucane HK, Ripke S, Yang J, Schizophrenia Working Group of the Psychiatric Genomics C, et al. LD Score regression distinguishes confounding from polygenicity in genome-wide association studies. Nature genetics. 2015;47(3):291–5.

83. McCarthy DJ, Campbell KR, Lun AT, Wills QF. Scater: pre-processing, quality control, normalization and visualization of single-cell RNA-seq data in R. Bioinformatics. 2017;33(8):1179–86.

84. Lun AT, McCarthy DJ, Marioni JC. A step-by-step workflow for low-level analysis of single-cell RNA-seq data with Bioconductor. F1000Res. 2016;5:2122.

85. Yu G, Wang LG, Han Y, He QY. clusterProfiler: an R package for comparing biological themes among gene clusters. Omics: a journal of integrative biology. 2012;16(5):284–7.

86. Burgess S, Foley CN, Allara E, Staley JR, Howson JMM. A robust and efficient method for Mendelian randomization with hundreds of genetic variants. Nature communications. 2020;11(1):376.

87. Burgess S, Small DS, Thompson SG. A review of instrumental variable estimators for Mendelian randomization. Statistical methods in medical research. 2017;26(5):2333–55.

88. Bowden J, Davey Smith G, Haycock PC, Burgess S. Consistent Estimation in Mendelian Randomization with Some Invalid Instruments Using a Weighted Median Estimator. Genetic epidemiology. 2016;40(4):304–14.

89. Burgess S, Thompson SG. Interpreting findings from Mendelian randomization using the MR-Egger method. European journal of epidemiology. 2017;32(5):377–89.

90. Verbanck M, Chen CY, Neale B, Do R. Detection of widespread horizontal pleiotropy in causal relationships inferred from Mendelian randomization between complex traits and diseases. Nature genetics. 2018;50(5):693–8.

91. Bowden J, Spiller W, Del Greco MF, Sheehan N, Thompson J, Minelli C, et al. Improving the visualization, interpretation and analysis of two-sample summary data Mendelian randomization via the Radial plot and Radial regression. Int J Epidemiol. 2018;47(4):1264–78.

92. Bowden J, Davey Smith G, Burgess S. Mendelian randomization with invalid instruments: effect estimation and bias detection through Egger regression. Int J Epidemiol. 2015;44(2):512–25.

93. Sanderson E, Davey Smith G, Windmeijer F, Bowden J. An examination of multivariable Mendelian randomization in the single-sample and two-sample summary data settings. Int J Epidemiol. 2019;48(3):713–27.

94. Wu P, Gifford A, Meng X, Li X, Campbell H, Varley T, et al. Mapping ICD-10 and ICD-10-CM Codes to Phecodes: Workflow Development and Initial Evaluation. JMIR Med Inform. 2019;7(4):e14325.

95. Bulik-Sullivan B, Finucane HK, Anttila V, Gusev A, Day FR, Loh PR, et al. An atlas of genetic correlations across human diseases and traits. Nature genetics. 2015;47(11):1236–41.

96. Shi H, Mancuso N, Spendlove S, Pasaniuc B. Local Genetic Correlation Gives Insights into the Shared Genetic Architecture of Complex Traits. American journal of human genetics. 2017;101(5):737–51.

97. Pickrell JK, Berisa T, Liu JZ, Segurel L, Tung JY, Hinds DA. Detection and interpretation of shared genetic influences on 42 human traits. Nature genetics. 2016;48(7):709–17.

98. Pickrell JK. Joint analysis of functional genomic data and genome-wide association studies of 18 human traits. American journal of human genetics. 2014;94(4):559–73.

99. Berisa T, Pickrell JK. Approximately independent linkage disequilibrium blocks in human populations. Bioinformatics. 2016;32(2):283–5.

