## Supplementary Figures for "GENOME-WIDE ASSOCIATIONS OF AORTIC DISTENSIBILITY SUGGEST CAUSAL RELATIONSHIPS WITH AORTIC ANEURYSMS AND BRAIN WHITE MATTER HYPERINTENSITIES"

**Supplementary Figure S1**

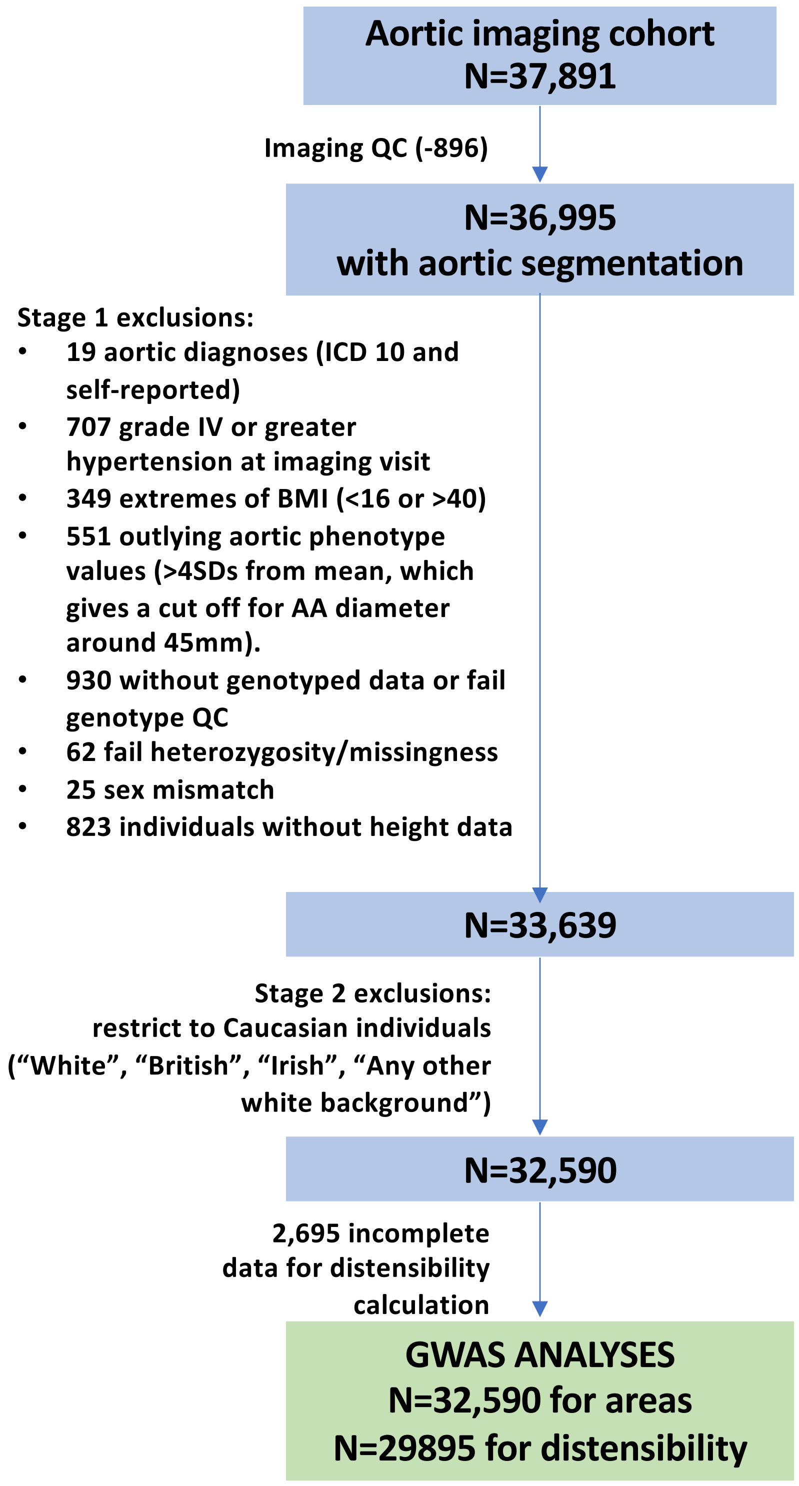


**Supplementary Figure S1: Details of exclusions**

#### Supplementary Figure S2

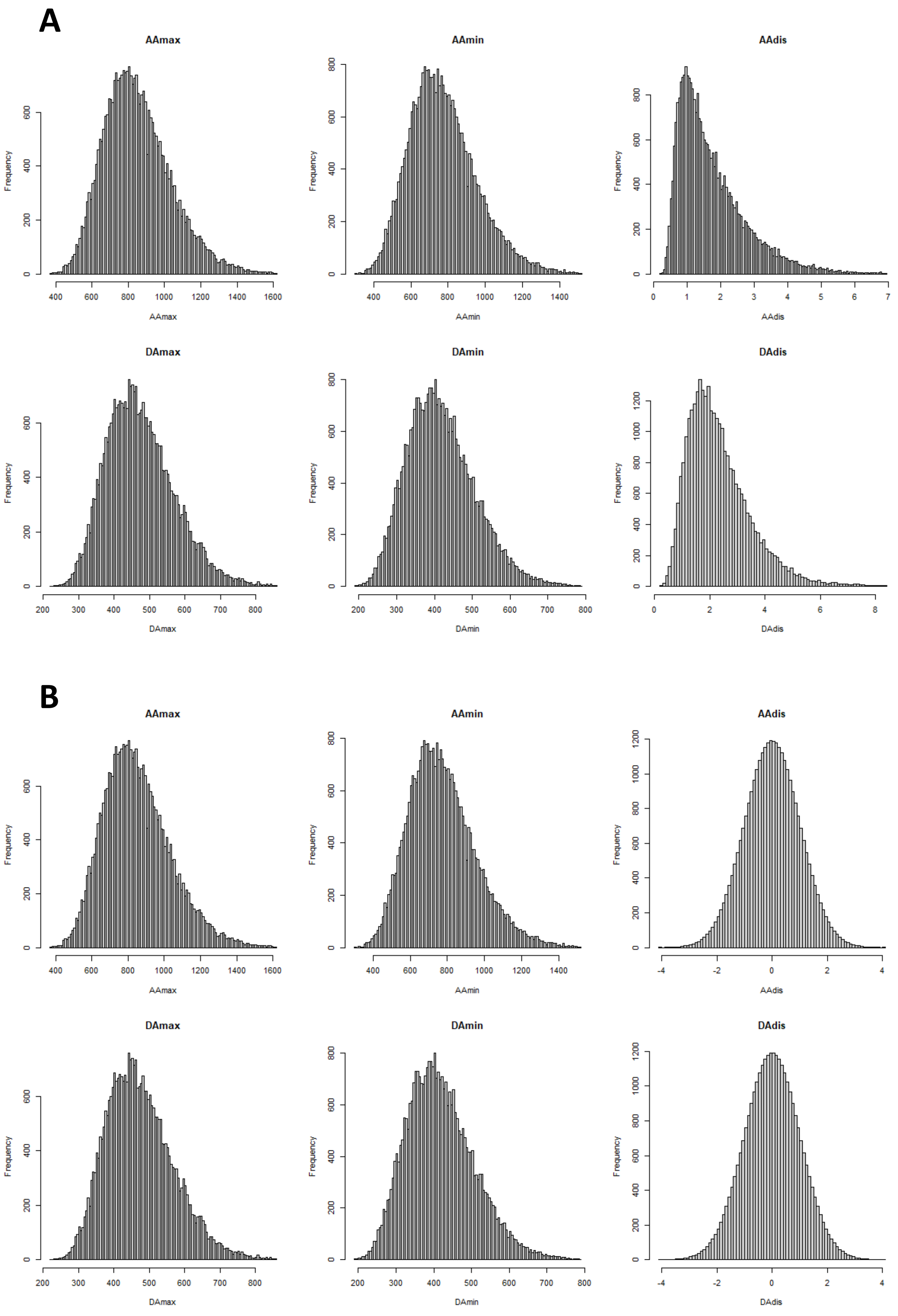


**Supplementary Figure S2: Distribution of image phenotype measures: A**) Original data and **B**) after rank normalisation for distensibility measures has been applied.

#### Supplementary Figure S3

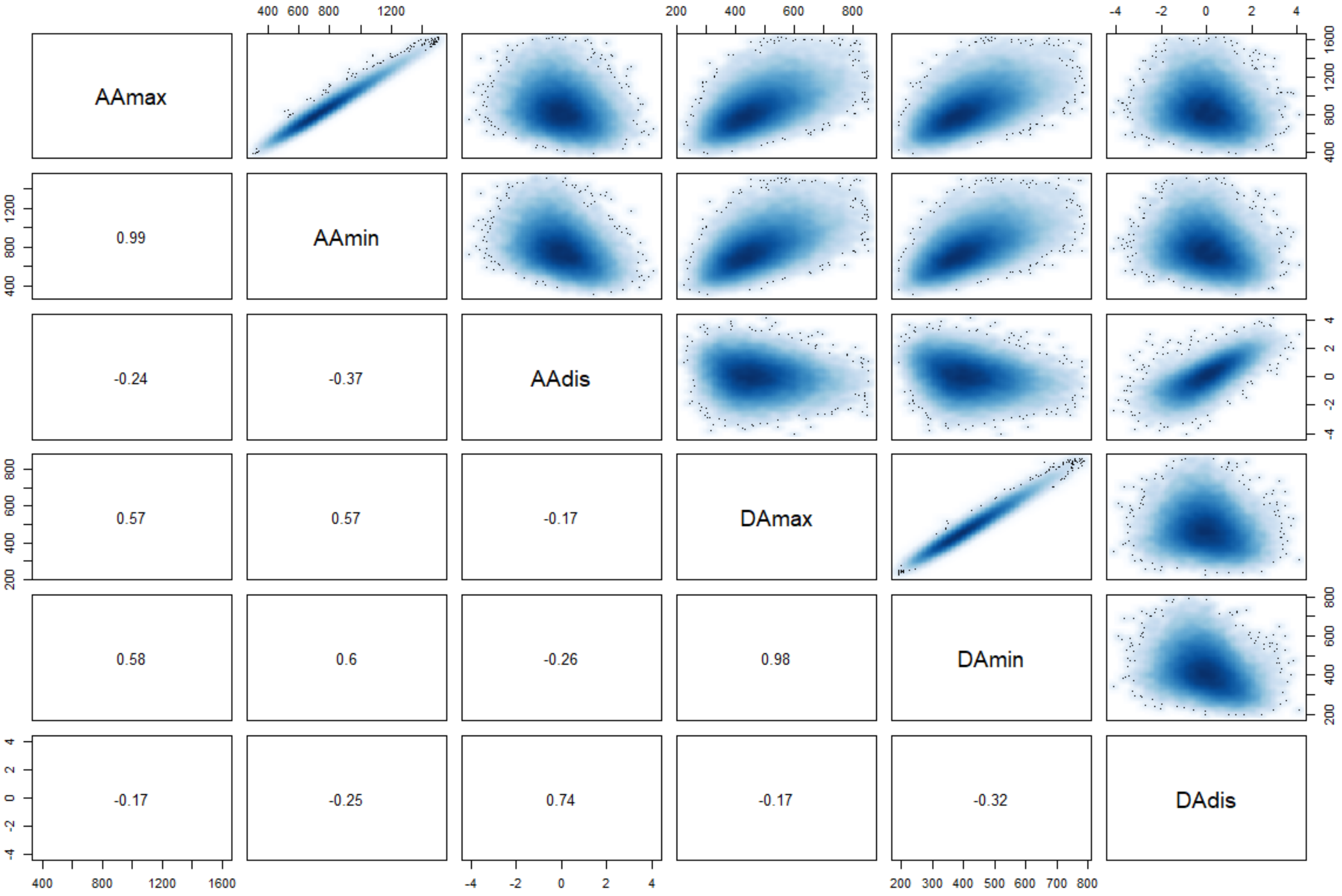


**Supplementary Figure S3: Correlation between imaging phenotypes**. Upper panels display smooth scatter plots, lower panels show the pair-wise Pearson's correlation coefficient. Plots or coefficients for distensibility phenotypes are based on rank normalised data. $p<2.2x10^{-16}$ for all comparisons.

#### Supplementary Figure S4

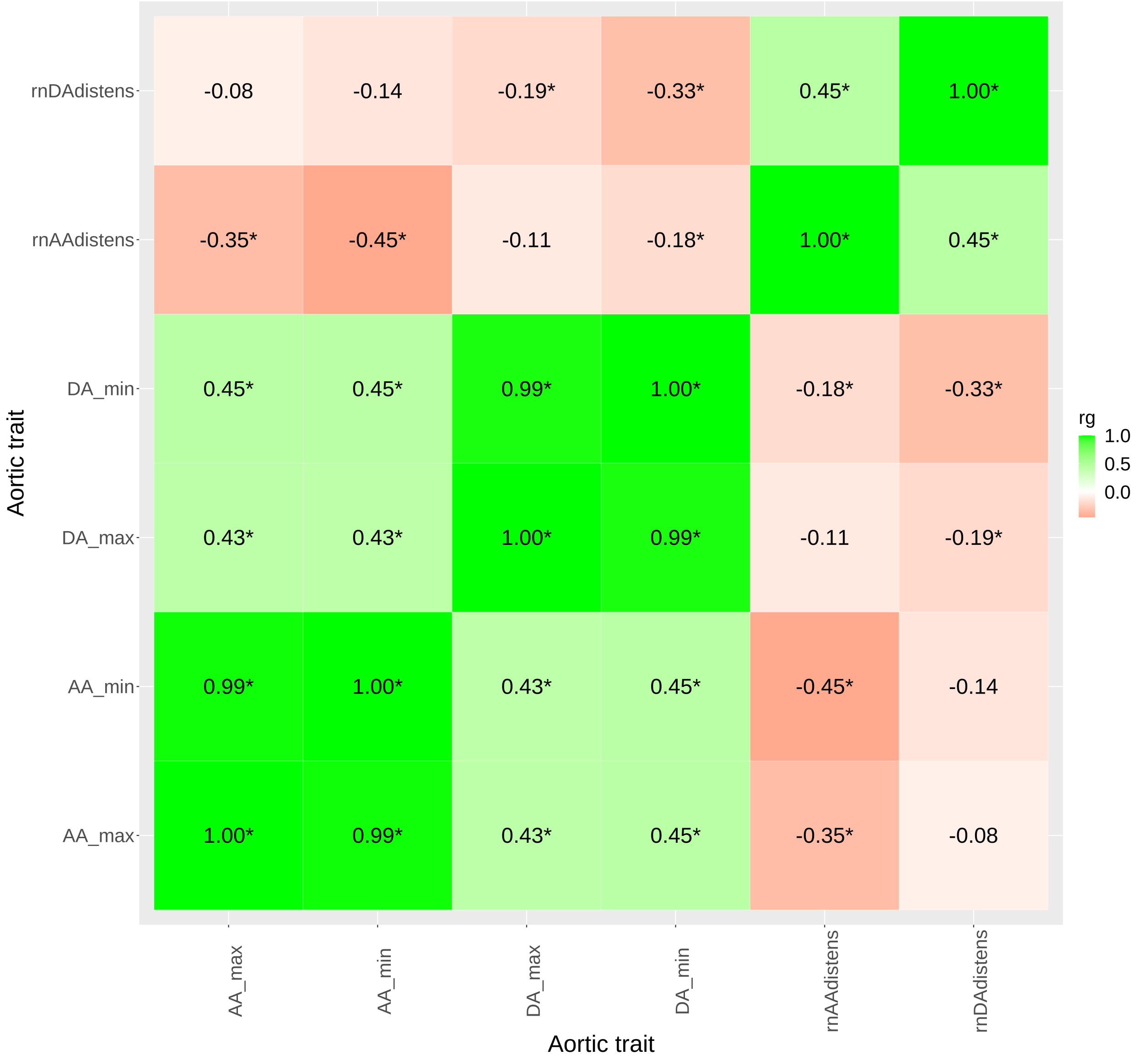


**Supplementary Figure S4. Genetic correlation between traits using LD score regression** (see Methods for details). Correlation coefficients are presented ($r_g$), with green representing positive correlation and red a negative correlation coefficient. AA_max: maximum ascending aortic area; AA_min: minimum ascending aortic area; DA_max: maximum descending aortic area; DA_min: minimum descending aortic area; rnAAdistens: rank-normalised ascending aortic distensibility; rnDAdistens: rank-normalised descending aortic distensibility. Asterisk indicates p value of correlation <0.05.

#### Supplementary Figures S5: Manhattan plots - Stage 1 GWAS

**A**

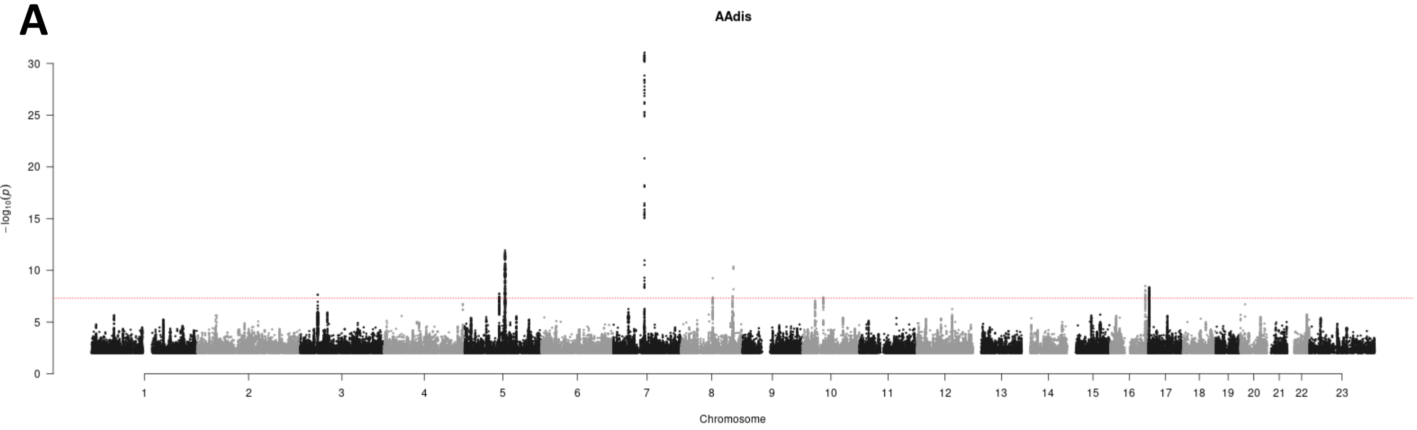


**B**

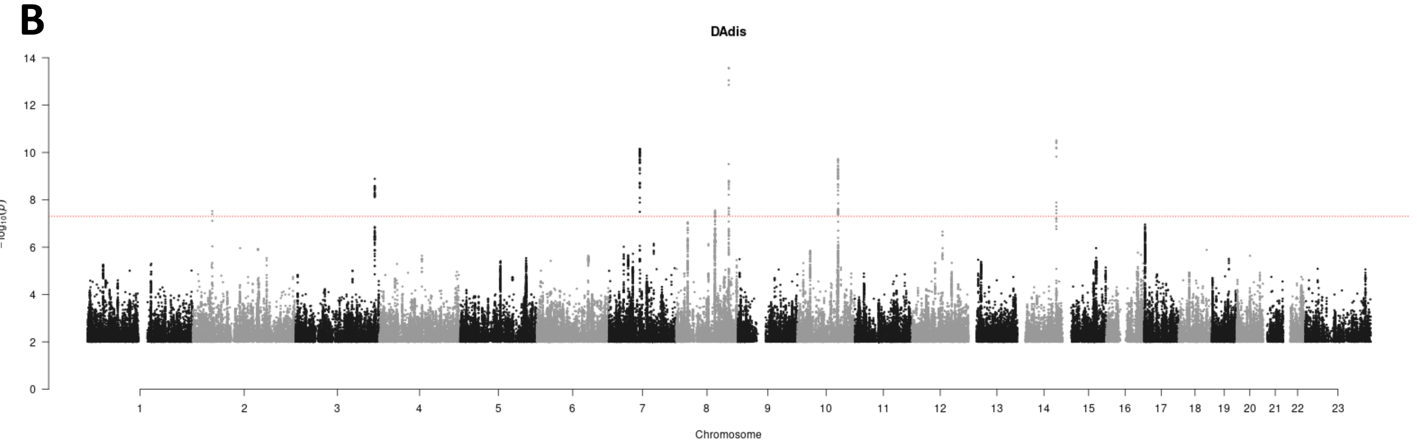


**C**

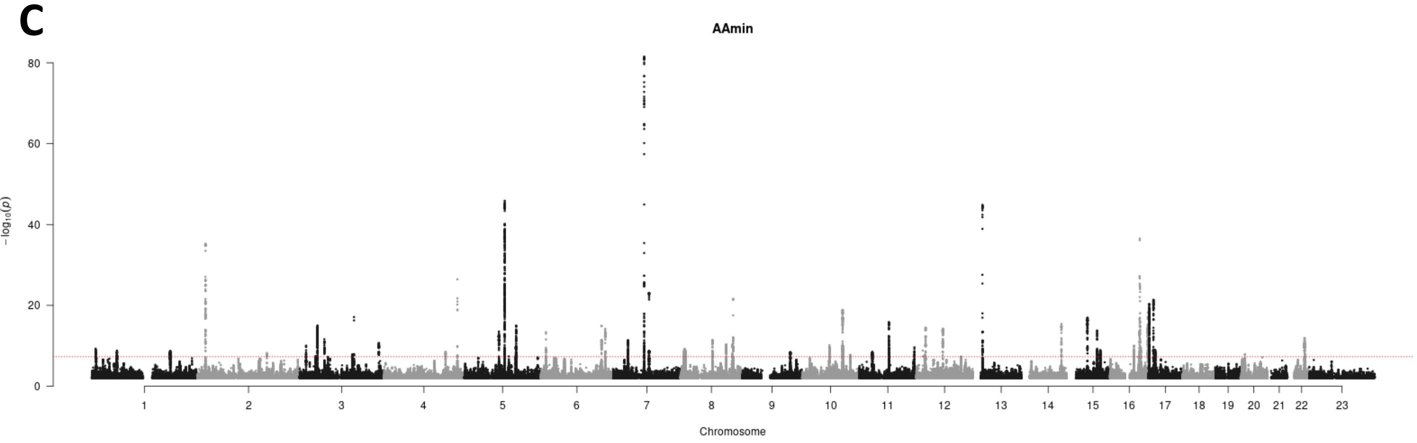


**D**

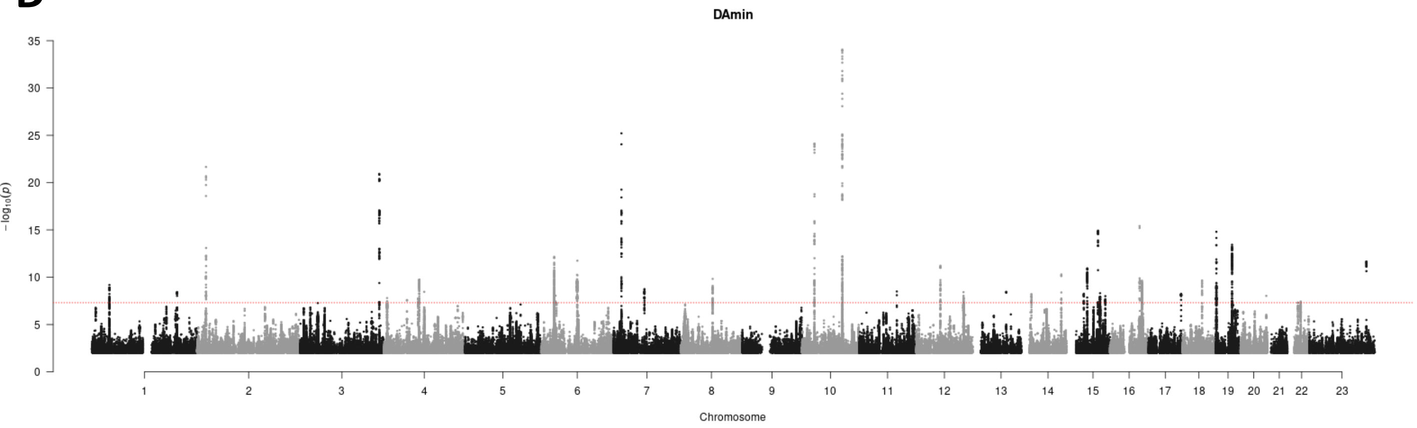

#### Supplementary Figures S5 cont…

E

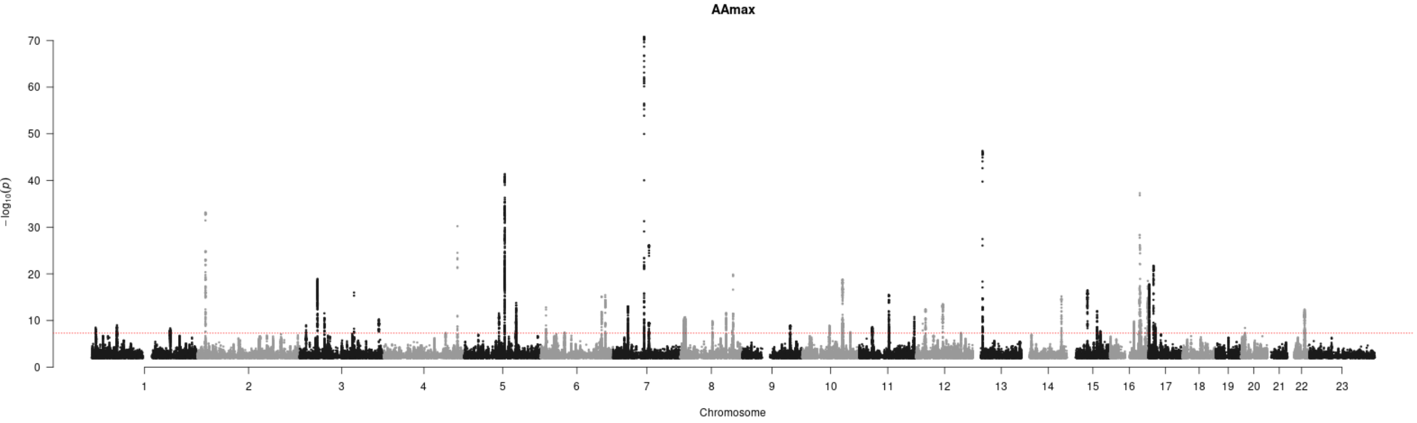


F

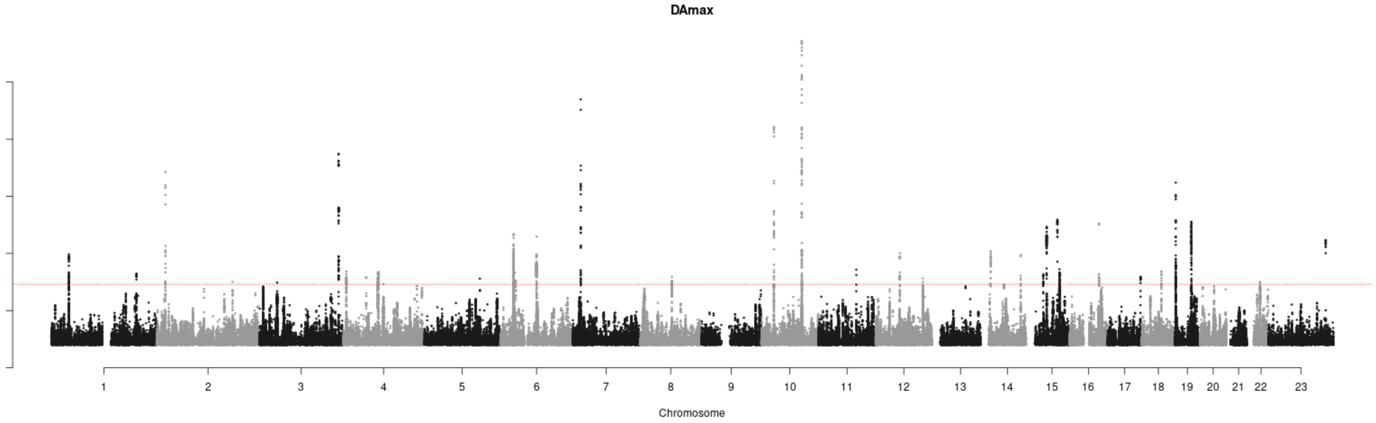


**Supplementary Figures S5:** Single-trait analysis results were obtained using BOLT-LMM. Summary statistics are shown as Manhattan plots with red dashed line showing the genome-wide significance threshold of $P = 5 \times 10^{-8}$. A= AAdis, B=DAdis, C=AAmin, D=DAmin, E=AAmax, F=DAmax.

#### Supplementary Figures S6: QQ plots - Stage 1 GWAS

AAdis

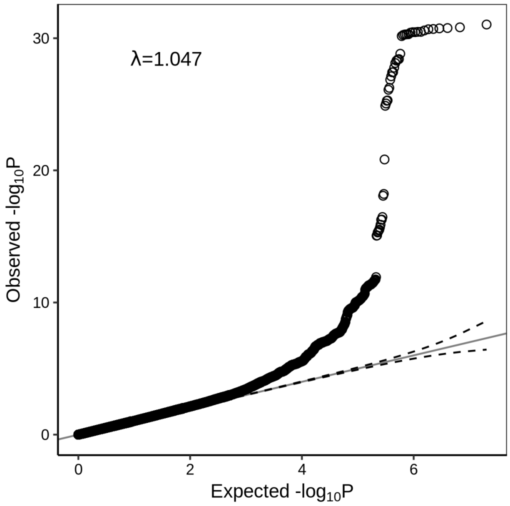


DAdis

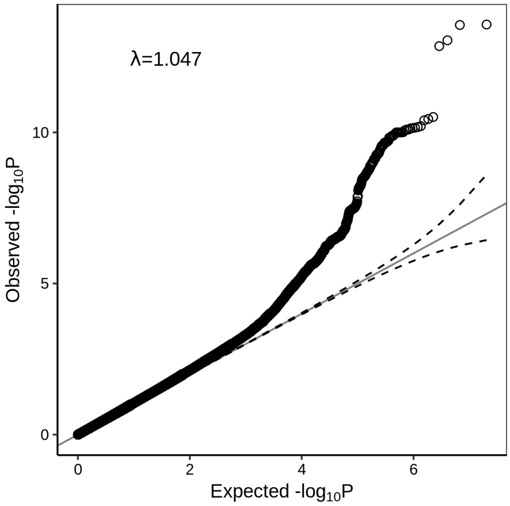


AAmin

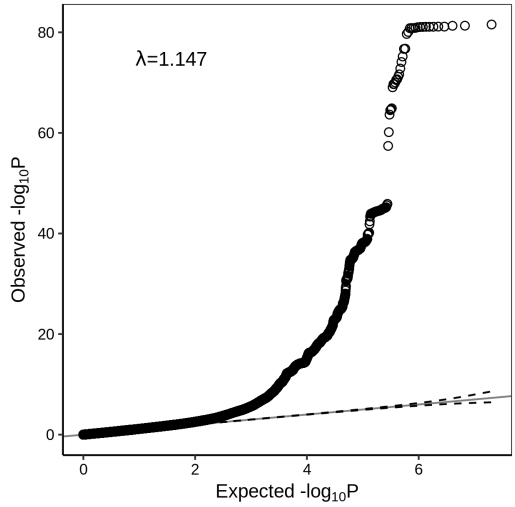


DAmin

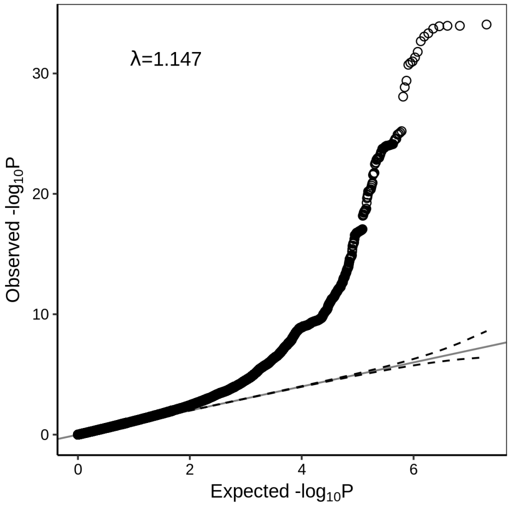


AAmax

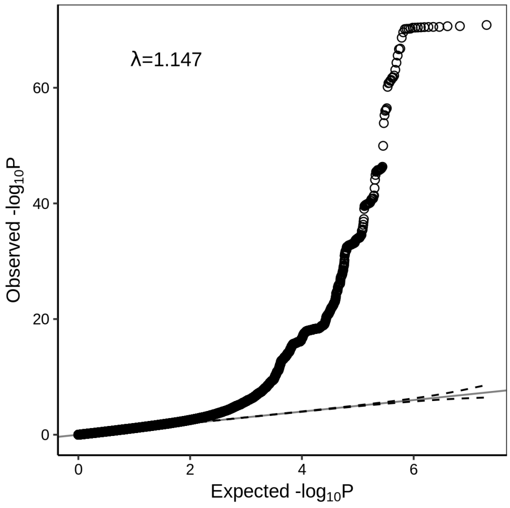


DAmax

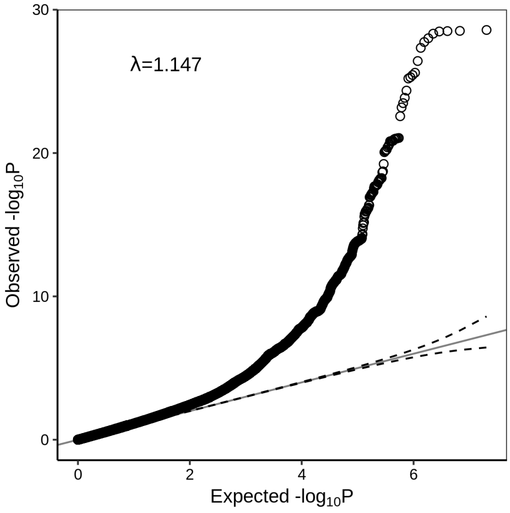


**Supplementary Figures S6: QQ plots showing observed versus expected $-\log_{10}$ p values for stage 1 GWAS, with genomic inflation factor in insert.**

### Supplementary Figures S7: Manhattan plots - Stage 2 (MTAG) GWAS

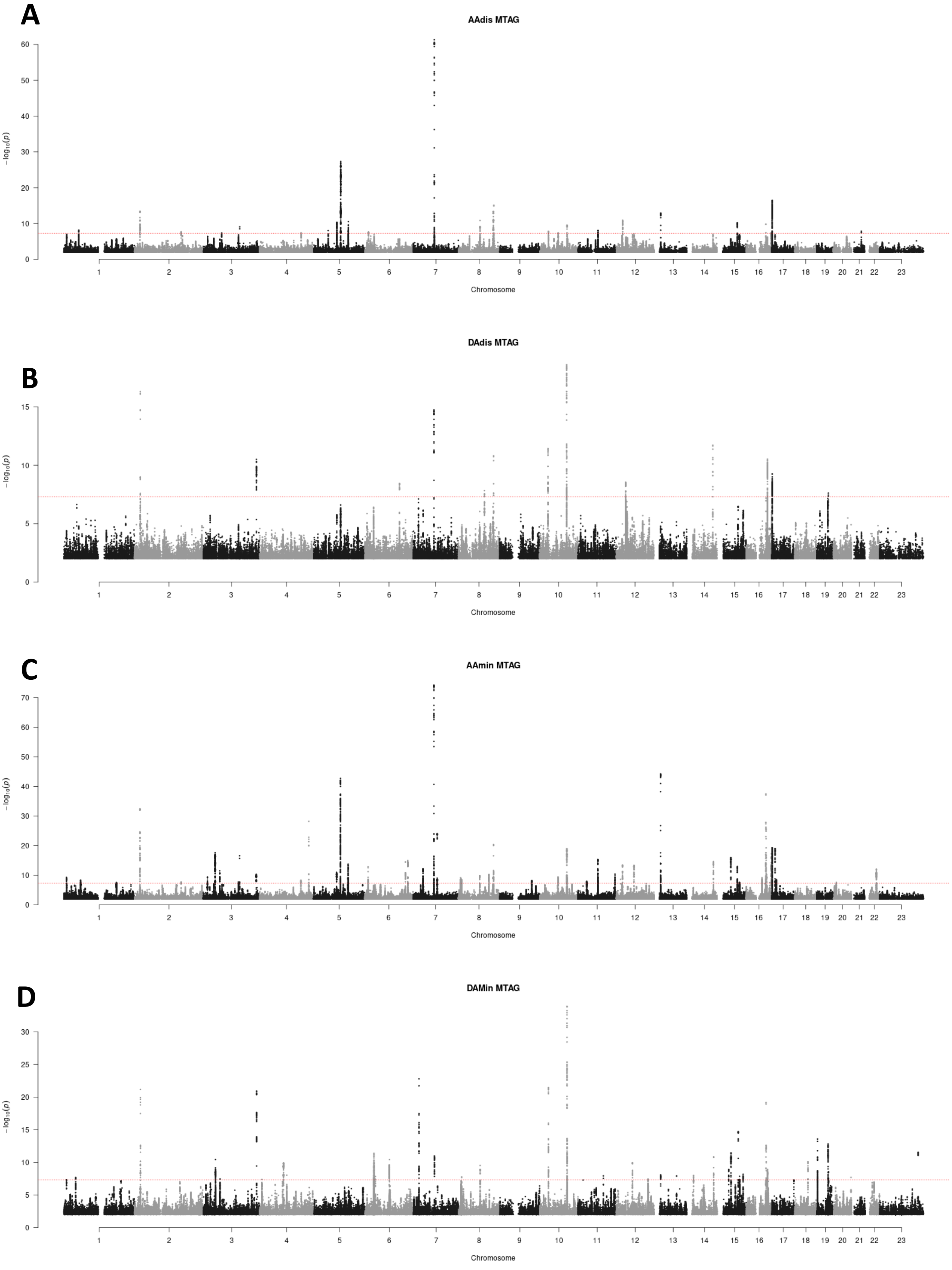

#### Supplementary Figures S7 cont…

E

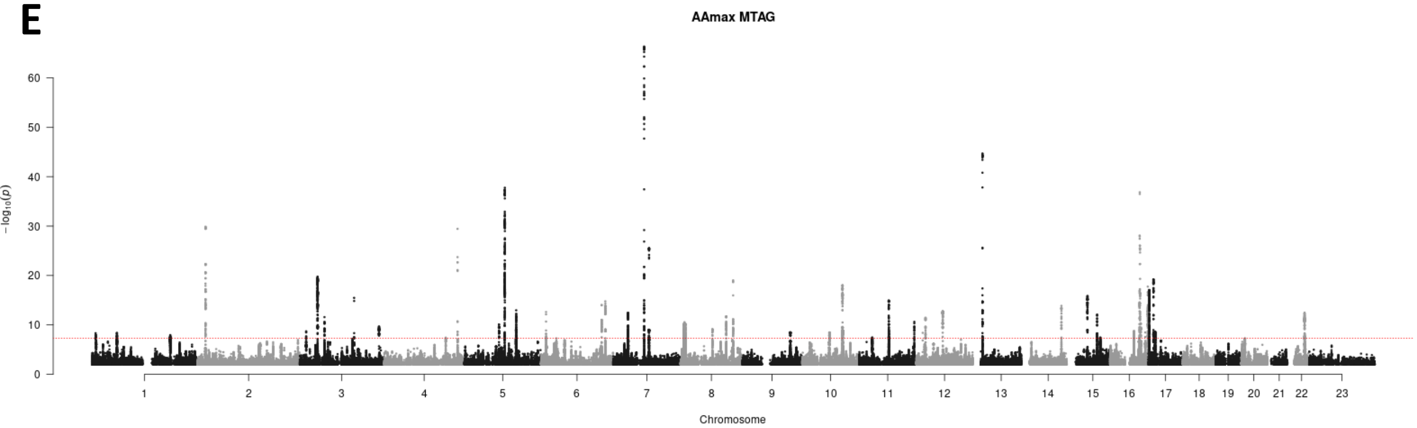


F

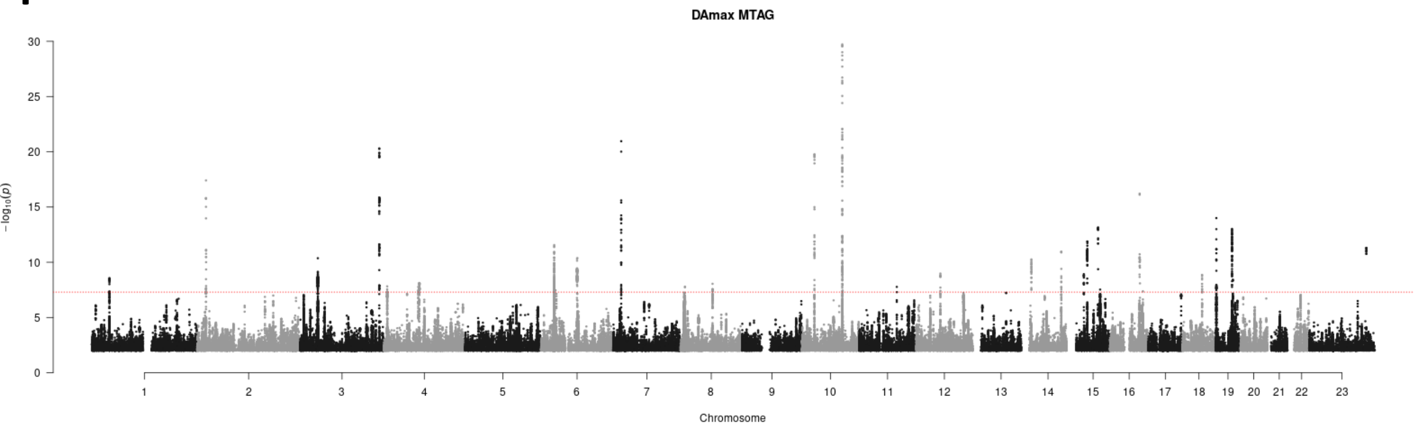


**Supplementary Figures S7 Manhattan plots showing multi-trait (MTAG) GWAS results using all six aortic traits.**. Summary statistics from MTAG are shown as Manhattan plots with red dashed line showing the genome-wide significance threshold of $P = 5 \times 10^{-8}$. A= AAdis, B=DAdis, C=AAmin, D=DAmin, E=AAmax, F=DAmax.

### Supplementary Figures S8

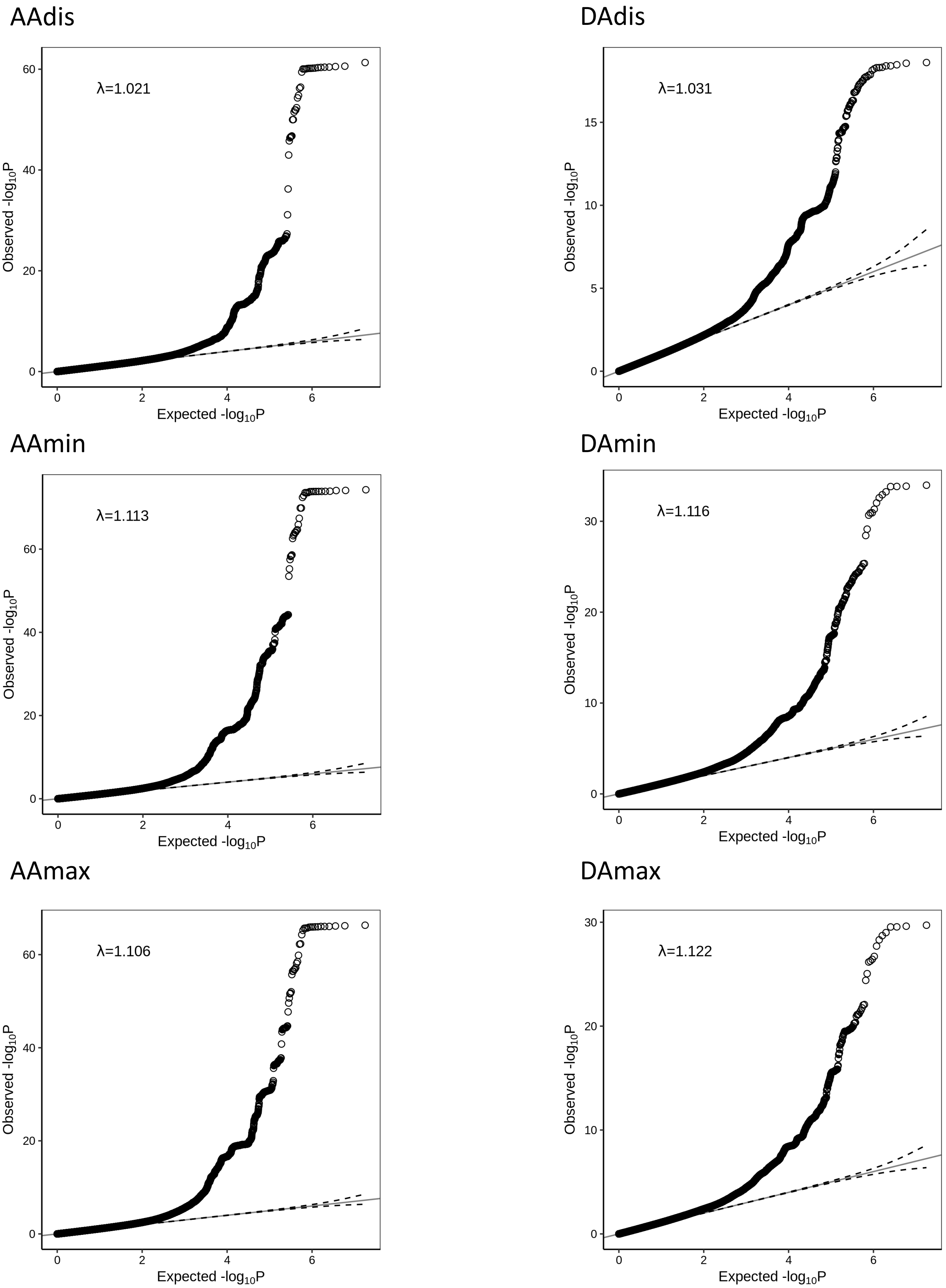


**Supplementary Figures S8: QQ plots showing observed versus expected -$\log_{10}$ p values for stage 2 (multi-trait) GWAS using MTAG, with genomic inflation factor (λ) in insert.**

#### Supplementary Figures S9

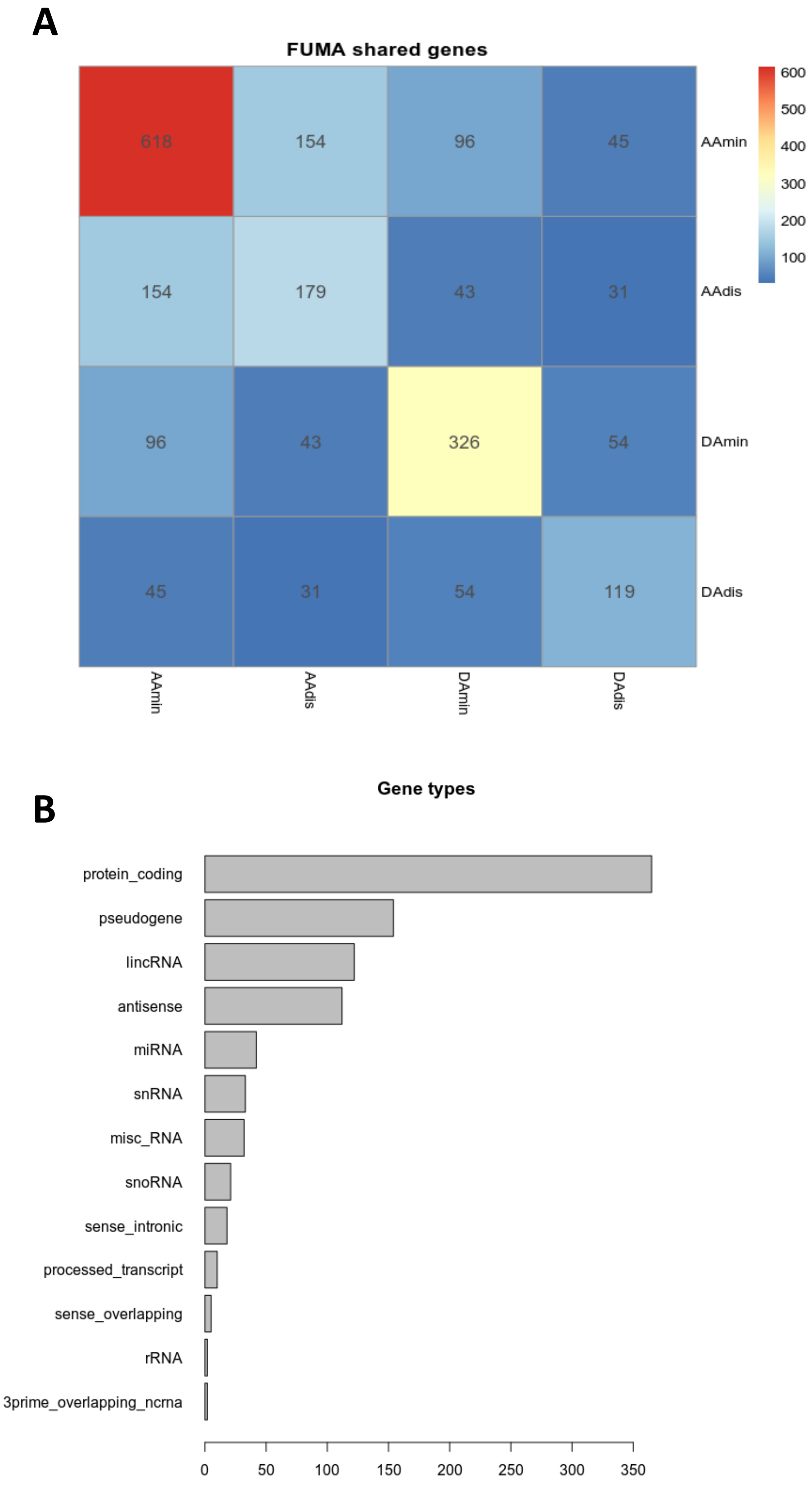


**Supplementary Figure S9: FUMA results:** (A) Overlap of genes detected for FUMA for different traits; (B) Gene types detected by FUMA

### Supplementary Figures S10

##### AAdis (MTAG)

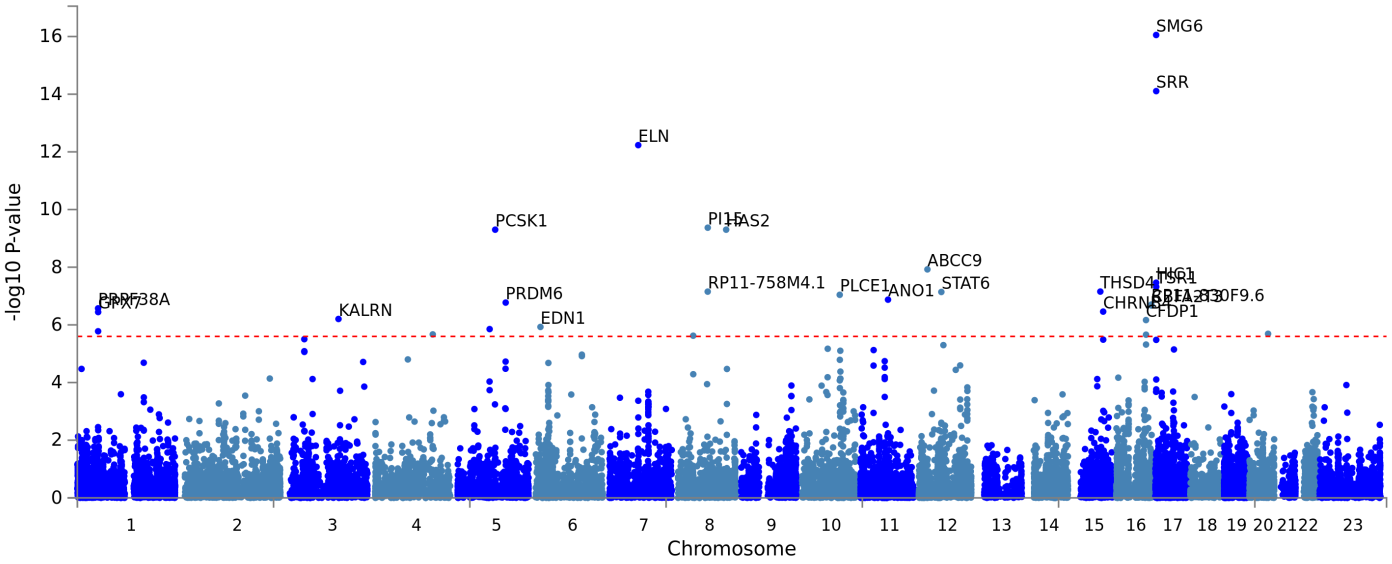


##### DAdis (MTAG)

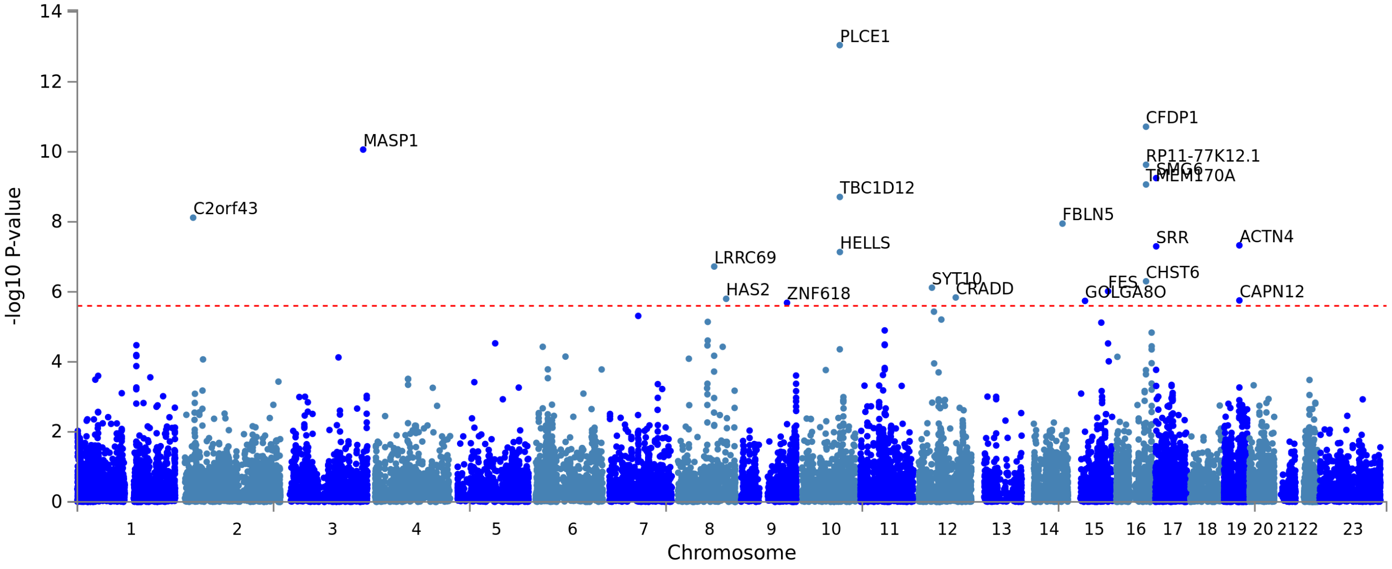


**Supplementary Figures S10: Manhattan plots showing gene-based analysis results from MAGMA (implemented in FUMA).** The top 21 genes associated with each trait are labelled.

#### Supplementary Figures S10 cont…

##### AAmin

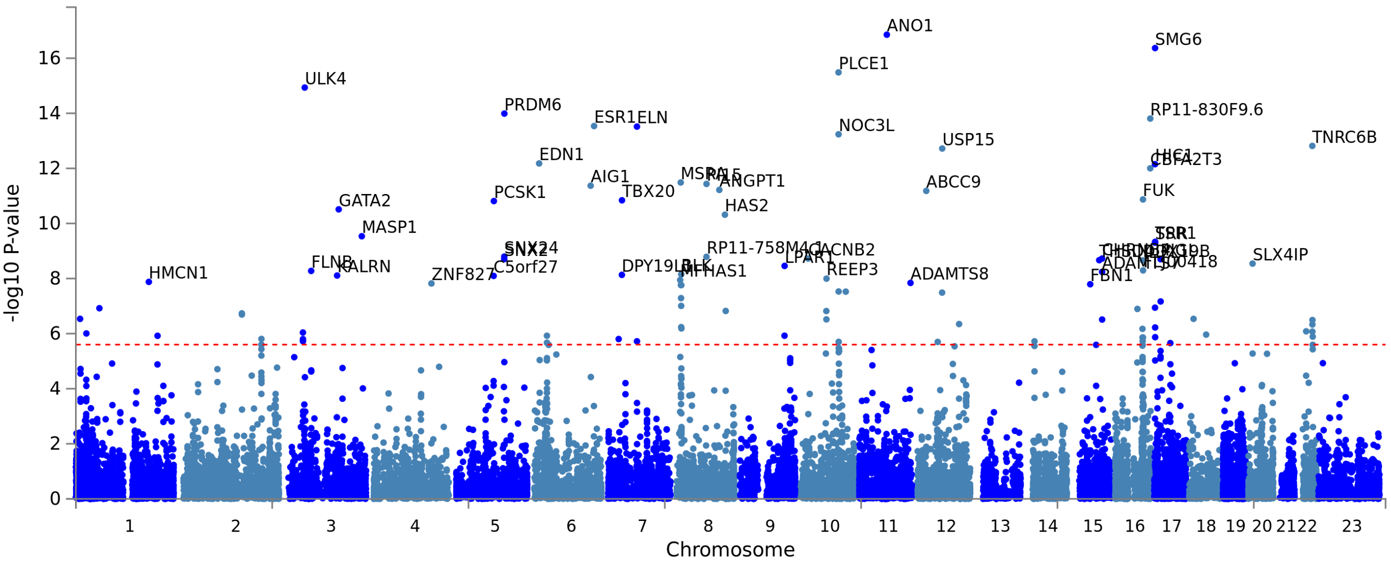


##### DAmin

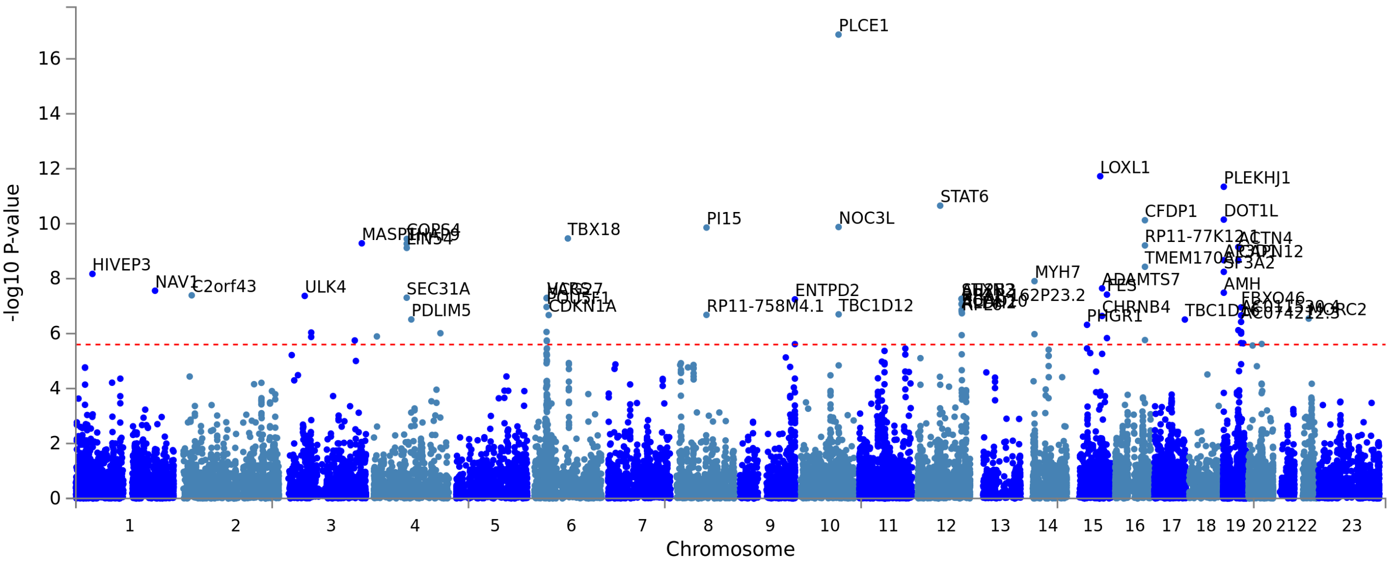


**Supplementary Figures S10 cont: Manhattan plots showing gene-based analysis results from MAGMA (implemented in FUMA).** The top 50 genes associated with each trait are labelled.

#### Supplementary Figure S11

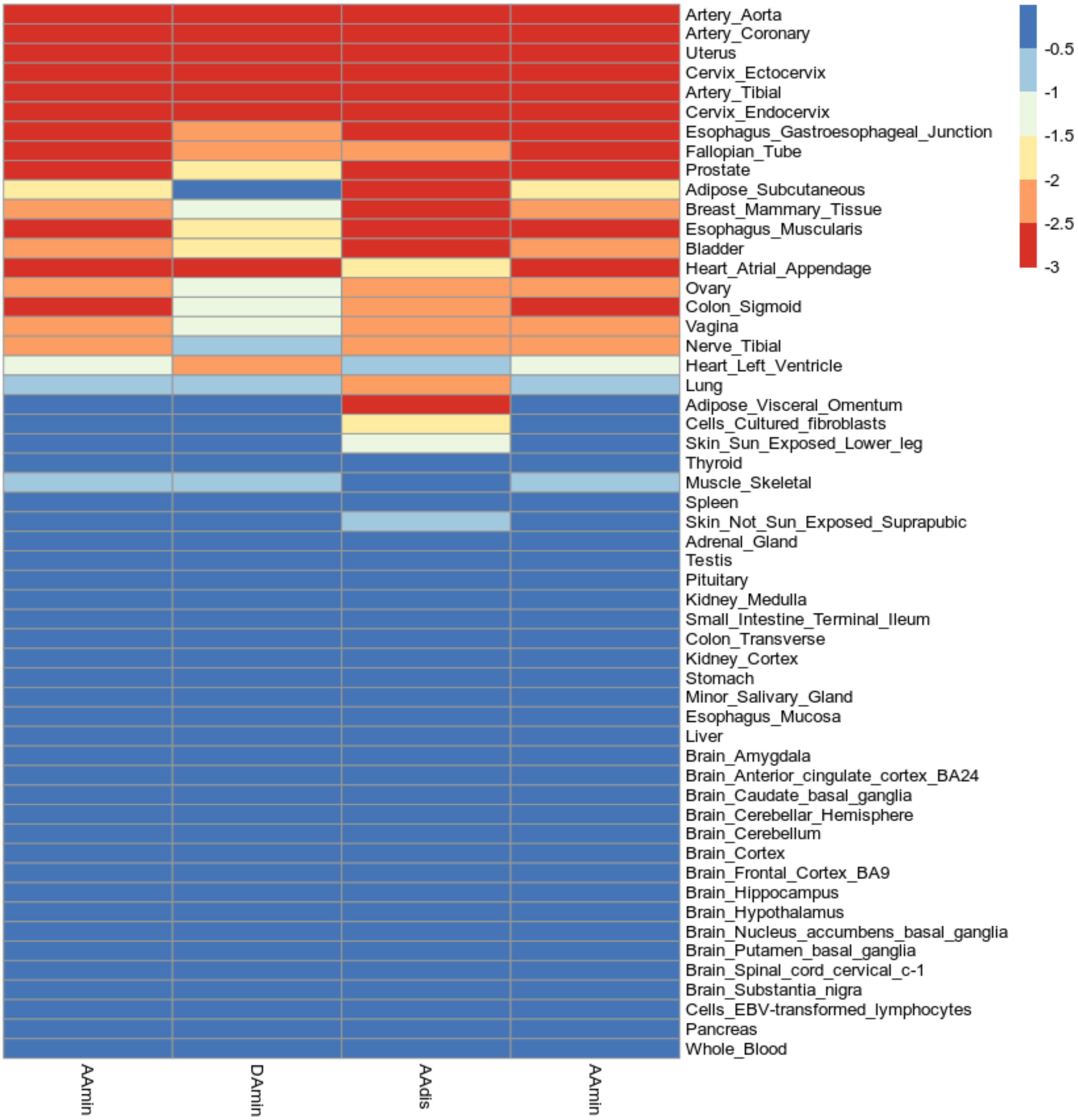


**Supplementary Figure S11: Heat map showing tissue expression analysis**, with the scale showing the –log10 p value of enrichment. Analysis in MAGMA, implemented in FUMA.

#### Supplementary Figure S12

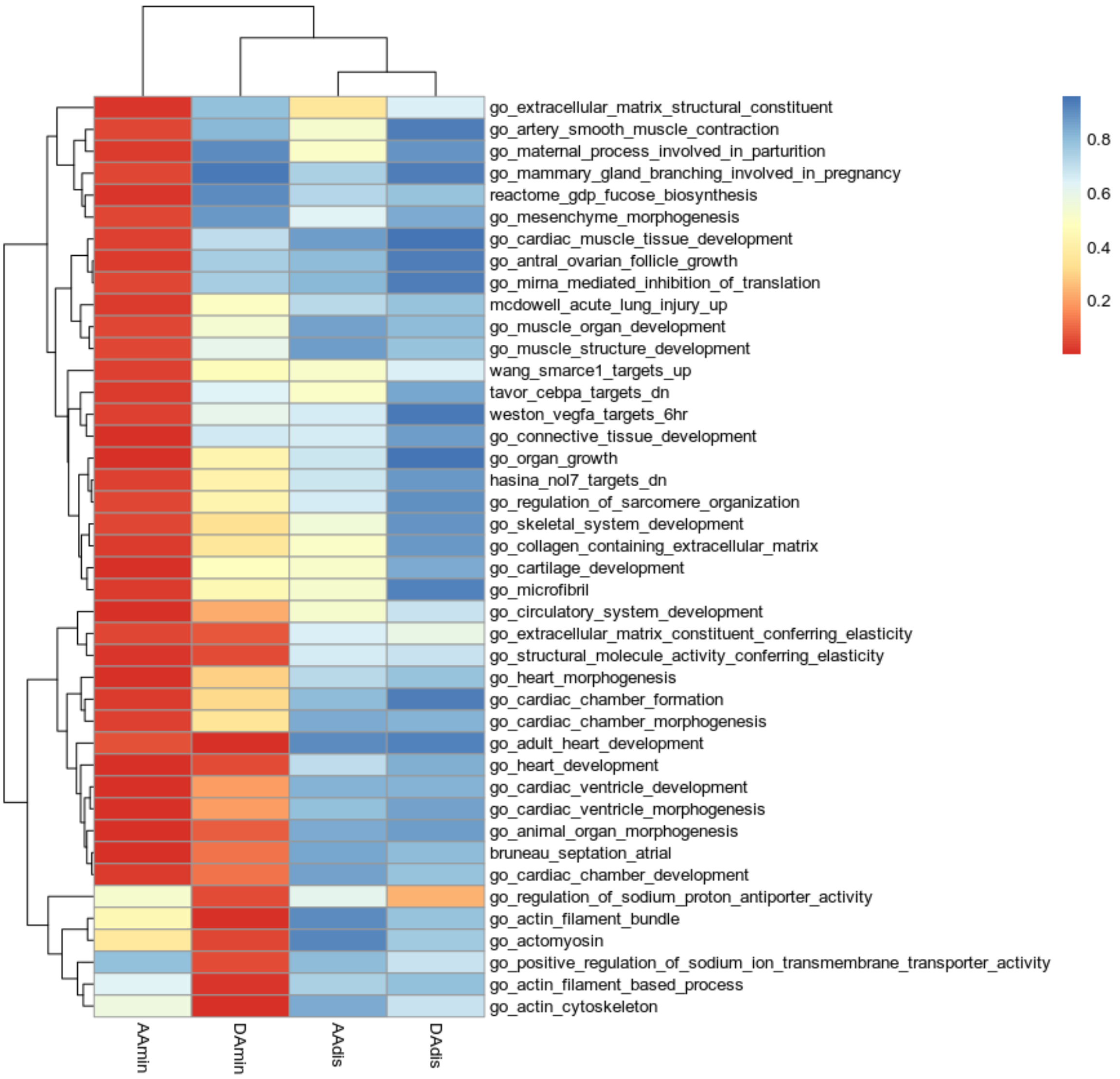


**Supplementary Figure S12: MAGMA gene set enrichment** Threshold FDR < 0.05 for at least one traits

#### Supplementary Figure S13

A

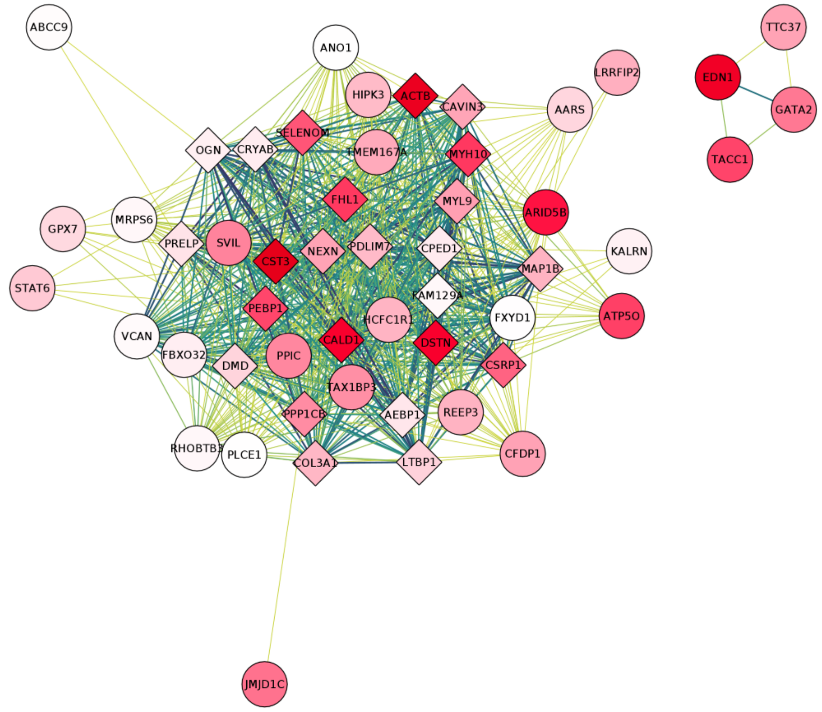


B

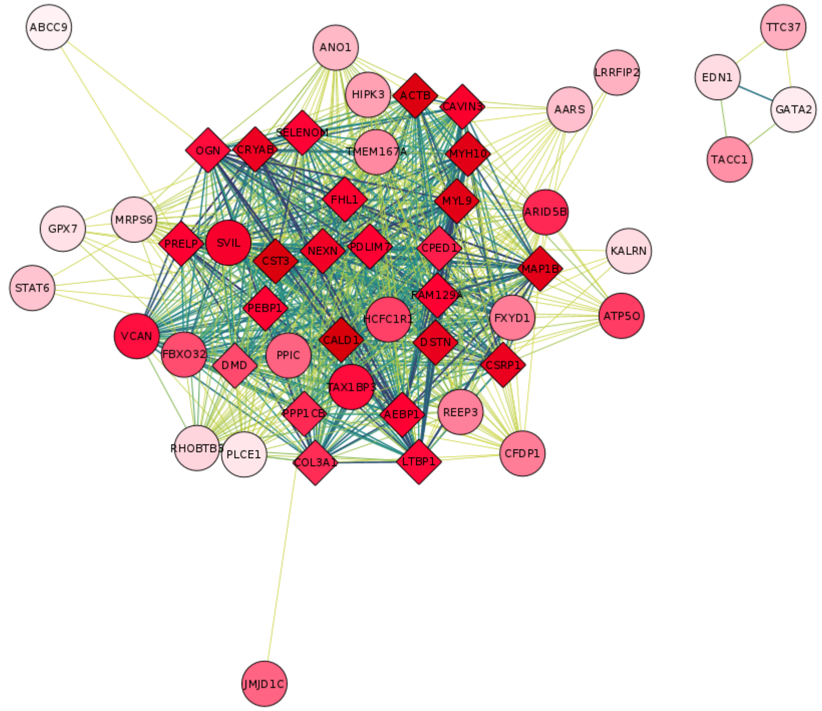


**Supplementary Figure S13: Extended coexpression networks: AAdis (MTAG)** Co-expression networks for aortic GWAS genes generated with primate single cell expression data for the aorta[33]. Shown are co-expression networks derived from extended models (r>0.1) in (A) endothelial and (B) smooth muscle cells. Round circles represent genes which were significantly associated with an aortic trait in the current GWAS. Diamonds represent other genes significantly co-expressed in the published single cell data for the cell-type indicated. The deeper the shade of red, the more highly that gene is expressed in the cell-type. The strength of co-expression is denoted by the colour of the lines joining genes with higher correlations indicated by darker lines. Hub genes are therefore found in the centres of these modules.

**Supplementary Figure S14**

A

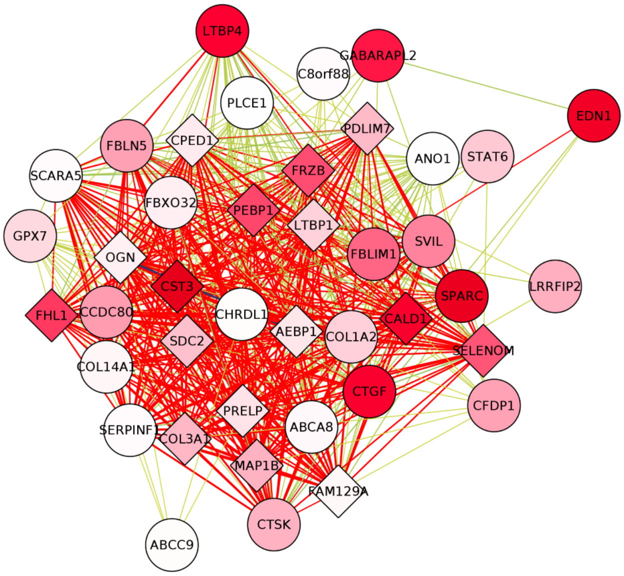


B

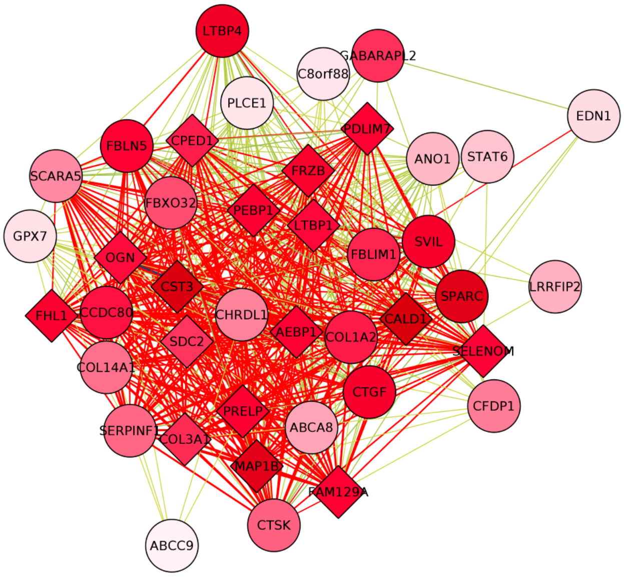


**Supplementary Figure S14: Extended coexpression networks: DAdis (MTAG).**Co-expression networks for aortic GWAS genes generated with primate single cell expression data for the aorta[33]. Shown are co-expression networks derived from extended models (r>0.1) in (A) endothelial and (B) smooth muscle cells. Round circles represent genes which were significantly associated with an aortic trait in the current GWAS. Diamonds represent other genes significantly co-expressed in the published single cell data for the cell-type indicated. The deeper the shade of red, the more highly that gene is expressed in the cell-type. The strength of co-expression is denoted by the colour of the lines joining genes with higher correlations indicated by darker lines. Hub genes are therefore found in the centres of these modules.

#### Supplementary Figure S15

A

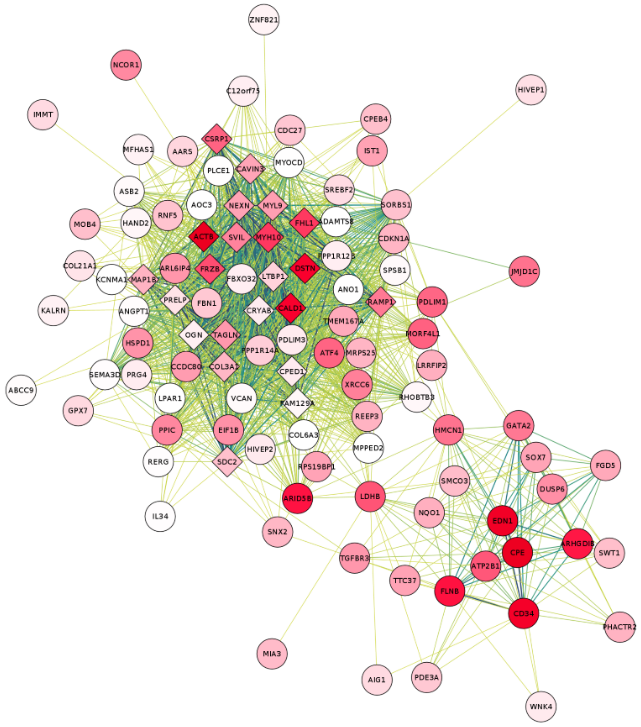


B

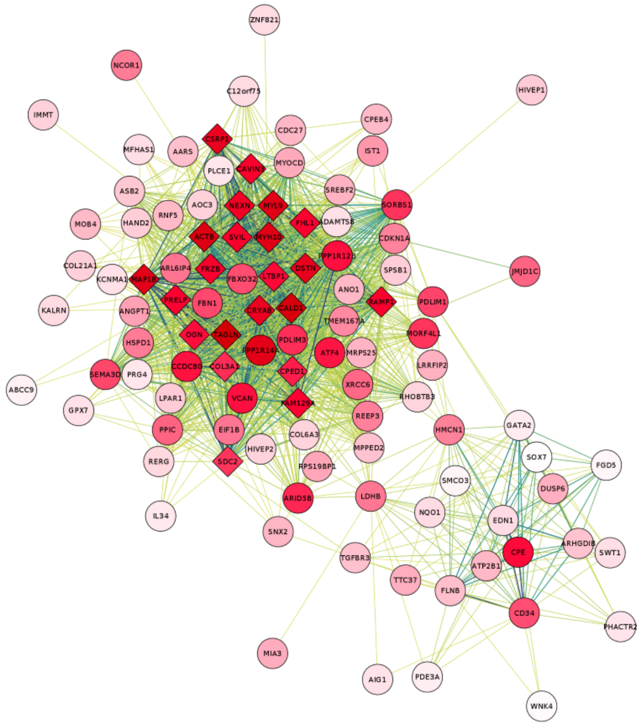


**Supplementary Figure S15: Extended coexpression networks: AAmin.** Co-expression networks for aortic GWAS genes generated with primate single cell expression data for the aorta[33]. Shown are co-expression networks derived from extended models (r>0.1) in (A) endothelial and (B) smooth muscle cells. Round circles represent genes which were significantly associated with an aortic trait in the current GWAS. Diamonds represent other genes significantly co-expressed in the published single cell data for the cell-type indicated. The deeper the shade of red, the more highly that gene is expressed in the cell-type. The strength of co-expression is denoted by the colour of the lines joining genes with higher correlations indicated by darker lines. Hub genes are therefore found in the centres of these modules.

#### Supplementary Figure S16

A

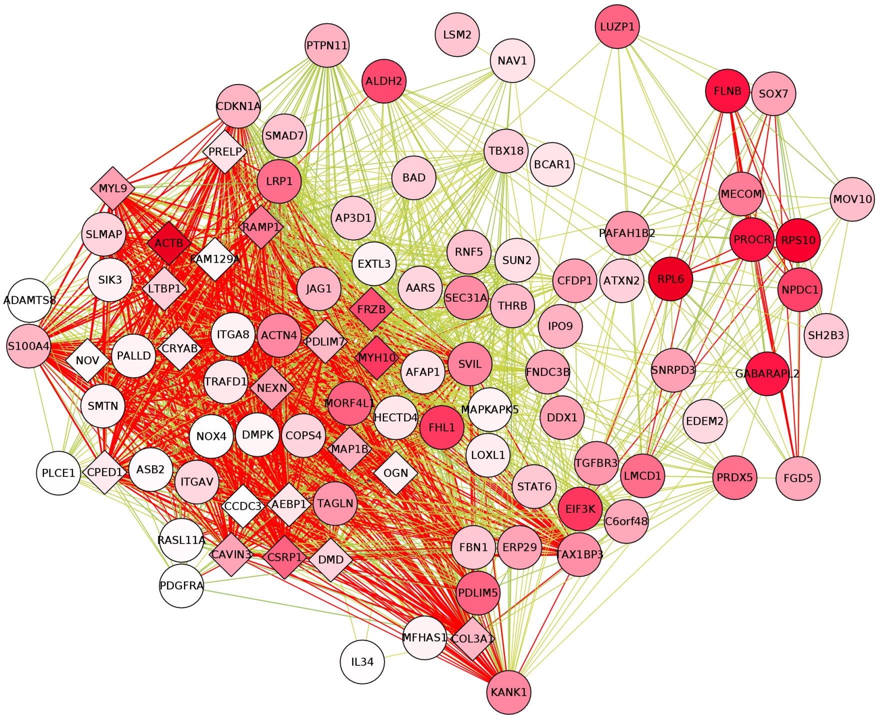


B

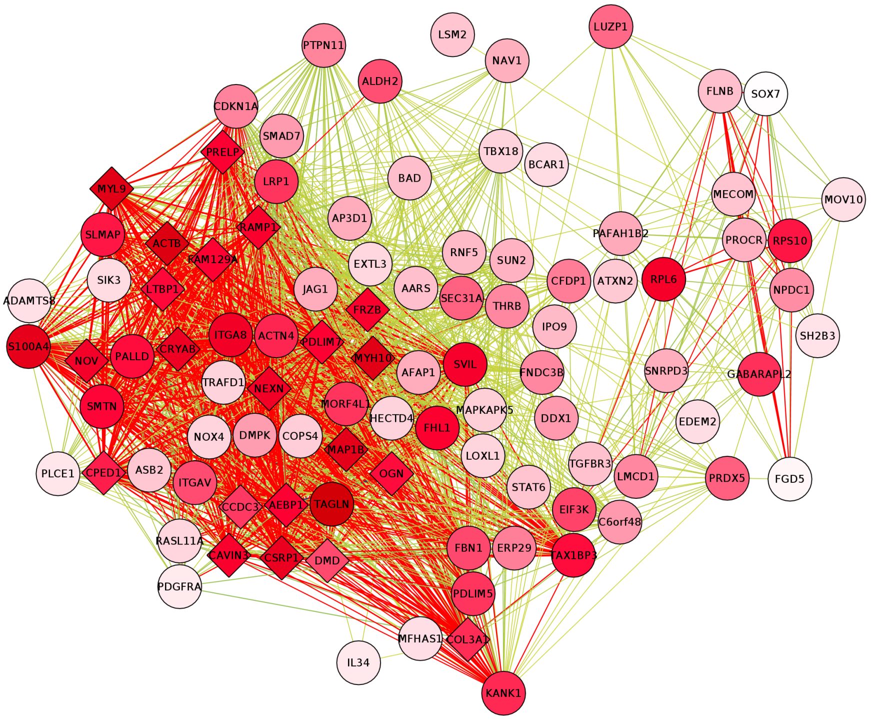


**Supplementary Figure S16: Extended coexpression networks: DAmin.** Co-expression networks for aortic GWAS genes generated with primate single cell expression data for the aorta[33]. Shown are co-expression networks derived from extended models (r>0.1) in (A) endothelial and (B) smooth muscle cells. Round circles represent genes which were significantly associated with an aortic trait in the current GWAS. Diamonds represent other genes significantly co-expressed in the published single cell data for the cell-type indicated. The deeper the shade of red, the more highly that gene is expressed in the cell-type. The strength of co-expression is denoted by the colour of the lines joining genes with higher correlations indicated by darker lines. Hub genes are therefore found in the centres of these modules.
